# The effect of livestock density on *Trypanosoma brucei gambiense* and *T. b. rhodesiense*: a causal inference-based approach

**DOI:** 10.1101/2022.01.13.22268995

**Authors:** Julianne Meisner, Agapitus Kato, Marshall Lemerani, Erick Mwamba Miaka, Acaga Ismail Taban, Jonathan Wakefield, Ali Rowhani-Rahbar, David Pigott, Jonathan Mayer, Peter Rabinowitz

**Affiliations:** Center for One Health Research, Department of Environmental and Occupational Health Sciences, University of Washington, Seattle, WA, USA; Department of Epidemiology, University of Washington, Seattle, WA, USA; Ministry of Agriculture, Animal Industry and Fisheries, Entebbe, Uganda; Ministry of Health, Lilongwe, Malawi; Programme National de Lutte contre la Trypanosomiase Humaine Africaine, Kinshasa, Democratic Republic of Congo; IntraHealth International, Juba, South Sudan; Department of Biostatistics, University of Washington, Seattle, WA, USA; Department of Statistics, University of Washington, Seattle, WA, USA; Department of Health Metrics Sciences, University of Washington, Seattle, WA, USA; Department of Global Health, University of Washington, Seattle, WA, USA

## Abstract

Domestic and wild animals are important reservoirs of the rhodesiense form of human African trypanosomiasis (rHAT), however quantification of this effect offers utility for deploying non-medical control activities, and anticipating their success when wildlife are excluded. Further, the uncertain role of animal reservoirs—particularly pigs—threatens elimination of transmission (EOT) targets set for the gambiense form (gHAT). Using a new time series of high-resolution cattle and pig density maps, HAT surveillance data collated by the WHO Atlas of HAT, and methods drawn from causal inference and spatial epidemiology, we conducted a retrospective ecological cohort study in Uganda, Malawi, Democratic Republic of Congo (DRC) and South Sudan to estimate the effect of cattle and pig density on HAT risk.

For rHAT, we found a positive effect for cattle (RR 1.61, 95% CI 0.90, 2.99) and pigs (RR 2.07, 95% CI 1.15, 2.75) in Uganda, and a negative effect for cattle (RR 0.88, 95% CI 0.71, 1.10) and pigs (RR 0.42, 95% CI 0.23, 0.67) in Malawi. For gHAT we found a negative effect for cattle in Uganda (RR 0.88, 95% CI 0.50, 1.77) and South Sudan (RR 0.63, 95% CI 0.54, 0.77) but a positive effect in DRC (1.17, 95% CI 1.04, 1.32). For pigs, we found a positive gHAT effect in both Uganda (RR 2.02, 95% CI 0.87, 3.94) and DRC (RR 1.23, 95% CI 1.10, 1.37), and a negative association in South Sudan (RR 0.66, 95% CI 0.50, 0.98).

While ecological bias may drive the findings in South Sudan, estimated E-values and simulation studies suggest unmeasured confounding and underreporting are unlikely to explain our findings in Malawi, Uganda, and DRC. Our results indicate cattle and pigs are important reservoirs of rHAT in Uganda but not Malawi, and that pigs—and possibly cattle–may be gHAT reservoirs.

**Author summary:** Domestic animals, including cattle and pigs, are known to be important for the transmission of the rhodesiense form of human African trypanosomiasis (rHAT), however the relative importance of these reservoirs compared to wild animals is uncertain and likely focus-specific. For the gambiense form (gHAT) transmission is predominantly human-to-human, however pigs are thought to be a possible reservoir. In this study we used pre-existing data on livestock density and HAT risk to estimate the strength of the effect of cattle and pig density on rHAT risk in Uganda and Malawi, and gHAT risk in Uganda, Democratic Republic of Congo (DRC), and South Sudan. We found evidence that cattle and pigs increase the risk of rHAT in Uganda but not Malawi, that pigs increase the risk of gHAT in Uganda and DRC, and that cattle increase the risk of gHAT in DRC alone. These results indicate that control of both forms of HAT should include domestic animals in a One Health framework, however control of rHAT in Malawi is unlikely to be achieved if measures exclude wild animals.

## Introduction

In recent decades, remarkable progress in the control of Human African trypanosomiasis (HAT)—a bloodborne protozoal parasite transmitted by the tsetse fly (*Glossina* species)—has led the WHO to set targets for elimination as public health problem (EPHP) by 2020, and elimination of transmission (EOT) by 2030. Global EPHP targets were met in 2018, however most endemic countries are not yet eligible for national EPHP validation, and there are significant challenges to achieving EOT goals [1].

Two forms of HAT, which are geographically- and epidemiologically-distinct, exist: the chronic form, caused by *Trypanosoma brucei gambiense* (gHAT) and endemic in western and central Africa, and the acute form, caused by *T. b. rhodesiense* (rHAT) and endemic in eastern and southern Africa. Both rHAT and gHAT present in two stages. The initial hemolymphatic stage, characterized by non-specific signs, is followed by the meningoencephalitic stage, characterized by central nervous system signs and eventual death in the absence of treatment, occurring within several weeks for rHAT and 18 months or longer for gHAT [2, 3].

Cattle are known to be an important reservoir of rHAT [4–8], and while pigs have historically been considered less important reservoirs due to their shorter lifespan, a survey conducted in Tanzania found 4.8% of domestic pigs harbored *T. b. rhodesiense* [9]. Numerous studies have also documented wildlife species to be competent rHAT reservoirs, and proximity to protected areas is a known risk factor for rHAT infection [10]. The zoonotic nature of rHAT complicates its control, leading to its exclusion from EOT goals [11] and significantly lower investment in rHAT surveillance and control compared with gHAT, raising concerns that rHAT will emerge as a major public health problem once gHAT EOT is achieved and donor attention moves away from HAT.

The omission of rHAT from EOT goals has been largely accepted by virtue of its low incidence, however rHAT is thought to be significantly underreported, due both to its acute presentation—limiting opportunities for diagnosis and utility of active screening activities—and absence of the priority status afforded to gHAT control and surveillance, with modeling work estimating up to 12 deaths occurred for every one reported death in an rHAT focus in Uganda [12]. In rHAT cases that are detected, detection is rarely in stage 1; with recent approval of a stage-independent oral therapeutic (fexinidazole) limited to gHAT [13], this leaves melarsoprol, a chemotherapeutic agent with an associated fatality of 5% [14], as the only treatment option. Finally, animal African trypanosomiasis (AAT) is an important limiting factor for livestock productivity and poverty alleviation in endemic areas [15], and non-medical control efforts for AAT and HAT coincide. The underreporting of and limited treatment options for rHAT, and the economic importance of AAT and opportunity to coordinate control of both diseases in a One Health framework, suggest that rHAT should not be overlooked.

With regards to gHAT, the Informal Expert Group on Gambiense HAT Reservoirs concluded in a recent review that latent human infections and animal reservoirs threaten EOT goals for gHAT, yet little is known of the role of animal reservoirs in the epidemiology of this form. The maintenance of certain HAT foci at hypo-endemic levels, the finding that human-derived *T. b. gambiense* strains cyclically transmitted between animals for more than a year remained human-infective [16], and the failure of some mathematical models to capture low prevalence dynamics without inclusion of an animal reservoir or invasion of infected tsetse, all suggest animal reservoirs for gHAT exist [11, 17–19]. Due to experimental evidence that *T. b. gambiense* can retain infectivity after repeated passage through pigs and that pigs may remain infected for prolonged durations (up to 18 months), the current consensus is that pigs are the animal reservoir most likely to threaten gHAT elimination [11, 16]. Furthermore, several important gHAT vectors have demonstrated a preference for pigs, who roam in humid and shady areas in the periphery of villages or along small rivers, resulting in high exposure to these riverine tsetse [20].

In this study, we use a time series of high-resolution cattle and pig density maps, HAT surveillance data collated by the WHO Atlas of HAT, and the parametric g-formula [21] to estimate the effect of livestock density on HAT risk. Defining livestock density as the ratio of animals to humans, we have conducted this analysis separately for each country, animal species, and gHAT and rHAT, in Malawi, Uganda, Democratic Republic of Congo (DRC), and South Sudan, four high-burden countries that do not yet meet eligibility criteria for EPHP validation. While there is ample evidence documenting the reservoir role of cattle in rHAT, the strength of this effect and the corresponding effect for pigs is likely focus-dependent. Country-specific estimates of the livestock effect, in concert with high-resolution data on livestock distribution, will provide both key parameters for rHAT modeling efforts and key inputs for national sleeping sickness control programs and other stakeholders wishing to identify sub-national foci where transmission may be going undetected, or where insecticide or trypanocide treatment of domestic animals may achieve EPHP (or even EOT). In gHAT foci, estimation of this effect will narrow critical knowledge gaps surrounding the role of domestic animals in gHAT transmission.

## Materials and methods

Our study is a retrospective ecological cohort study, with cluster-year (0.017° pixel) as the unit of analysis in Malawi, Uganda, and DRC, and county (administrative level 2) as the unit of analysis in South Sudan.

### Study population

In Uganda, DRC, and Malawi, our study population included all clusters within five hours’ travel time of a fixed health facility capable of HAT diagnosis [22]. In Uganda, where gHAT and rHAT coexist, study areas were defined separately for each form; only gHAT is endemic in DRC, and only rHAT is endemic in Malawi. In South Sudan, where gHAT is endemic, we restricted the study area to counties which reported one or more HAT cases or conducted active surveillance during the study period.

The study period was defined separately for each country, on the basis of WHO Atlas of HAT data access provided to the authors and available exposure data: 2006-2018 for Uganda, 2003-2014 for Malawi, and 2010-2013 for DRC. In South Sudan the study period was restricted to 2008 alone due to limited exposure data availability, as detailed below.

### DAG formulation

We identified confounders by a priori subject knowledge, which we encoded in a directed acyclic graph (DAG). We used an initial time-stratified DAG (Fig 1) and DAGitty.net [23] to identify the minimally sufficient adjustment set.

**Fig 1.**
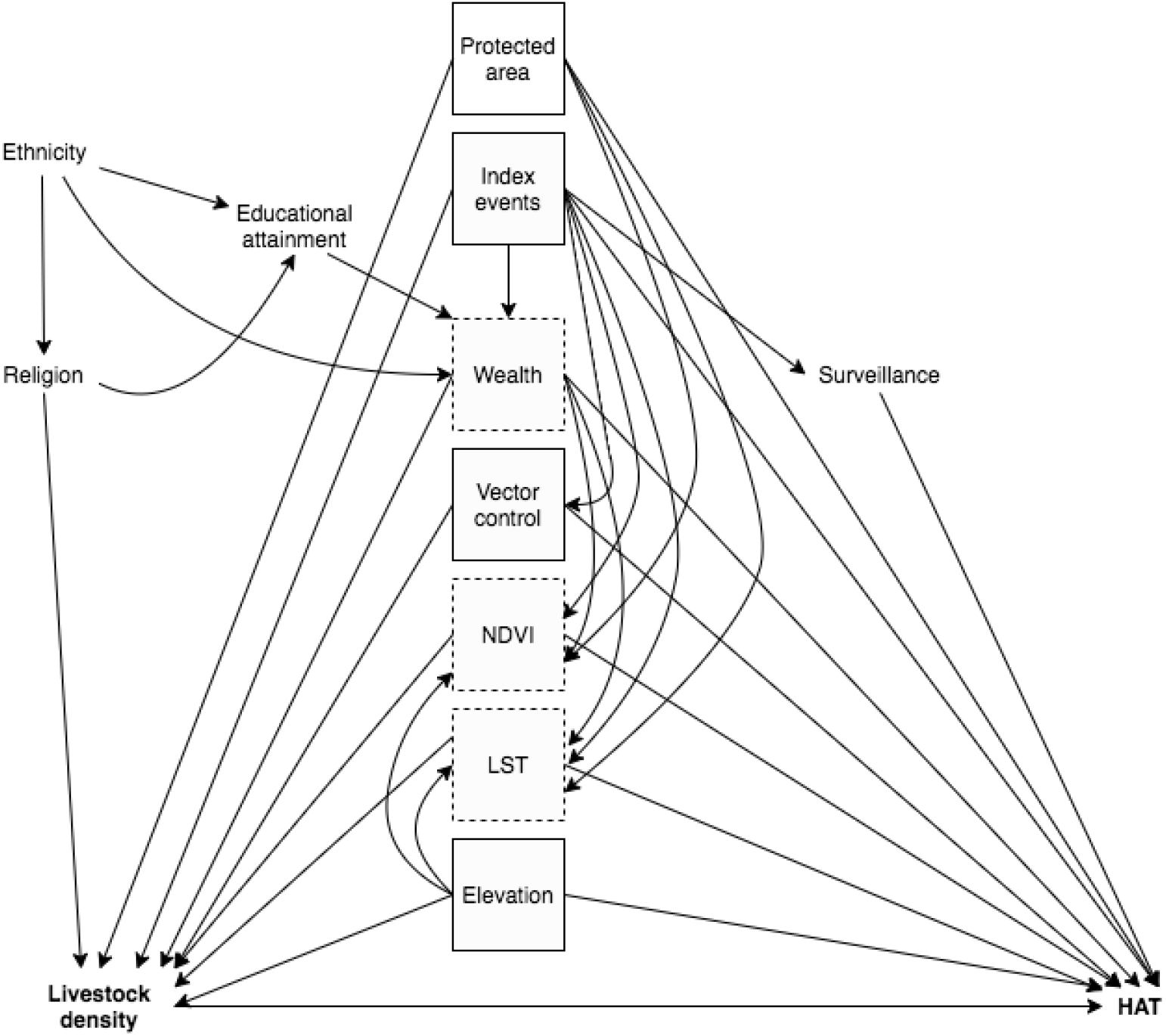
Time-stratified directed acyclic graph (DAG). Boxed variables are those in the minimum sufficient adjustment set; those with a dashed line exhibit exposure-confounder feedback. Bolded variables are the exposure and outcome of interest. Protected areas refers to rHAT models only.

“Index events” refers to natural and technological disasters and armed conflict. Location within a protected area was considered for rHAT alone, as a proxy for presence of wildlife reservoirs. Note that all variables are cluster-level, representing proportions for variables which are composites of individual-level categorical variables (ethnicity, educational attainment, and religion), and means for composites of continuous variables (wealth). The minimally sufficient adjustment set is {elevation, index events, LST, NDVI, wealth, vector control}.

The variables marked with a dashed box in Fig 1 exhibit exposure-confounder feedback: they are time-varying variables which are causes of the outcome and are both downstream of earlier exposure (mediators) and upstream of later exposure (confounders) (Fig 2). As unbiased estimation of the total exposure-outcome effect requires adjustment of confounders but precludes adjustment of mediators, traditional regression adjustment cannot be utilized. Robin’s generalized methods, referred to as “g-methods,” allow the identification and estimation of causal effects in the face of time-varying confounding with exposure-confounder feedback. As we do not have longitudinal exposure data in South Sudan, time-varying confounding is not a concern.

**Fig 2.**
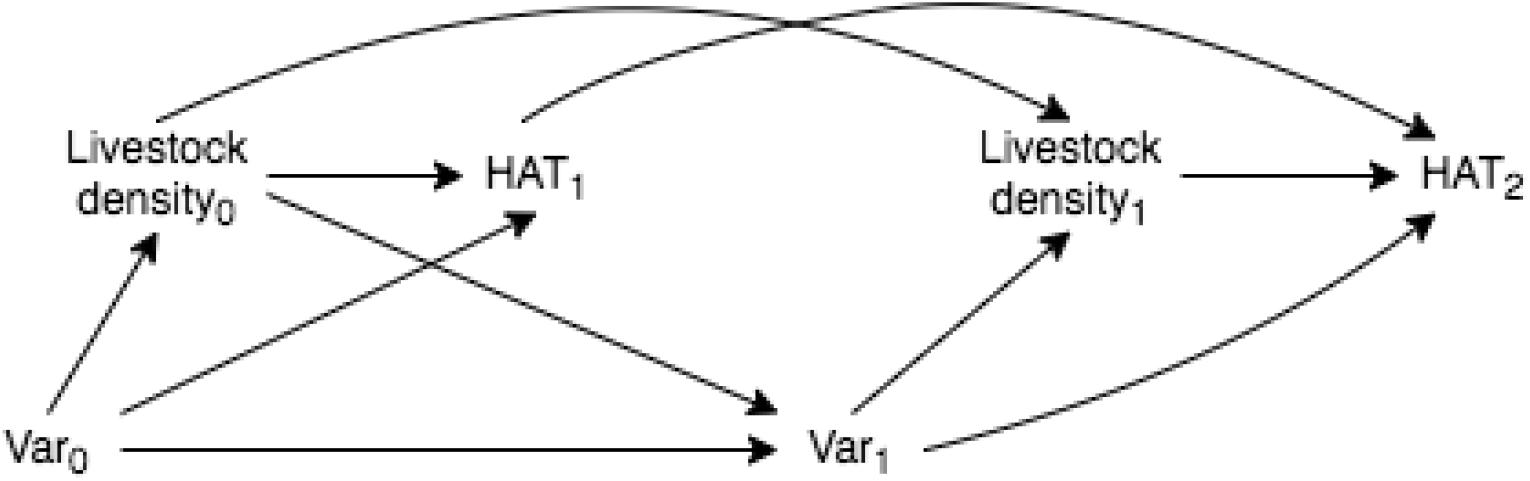
Time-varying directed acyclic graph (DAG) demonstrating exposure-confounder feedback. Var_0_ is a vector containing wealth, NDVI, and LST at the first of two hypothetical time points, and Var_1_ is a vector containing the same variables at the second time point. Var_1_ is a confounder of the livestock density_1_—HAT_2_ pathway, but is a mediator of the livestock density_0_—HAT_1_ and livestock density_0_—HAT_2_ pathways, thus adjustment of Var={Var_0_, Var_1_} will bias the joint (over time) effect of livestock density on HAT

Our final longitudinal DAG, restricted to the minimally sufficient adjustment set and to three time points, is presented in Fig 3. We assume index events are upstream of environmental variables (LST and NDVI) and wealth measured at the same time, but impose lags of one year for the effects of index events, environmental variables, and wealth on both livestock density and HAT risk.

**Fig 3.**
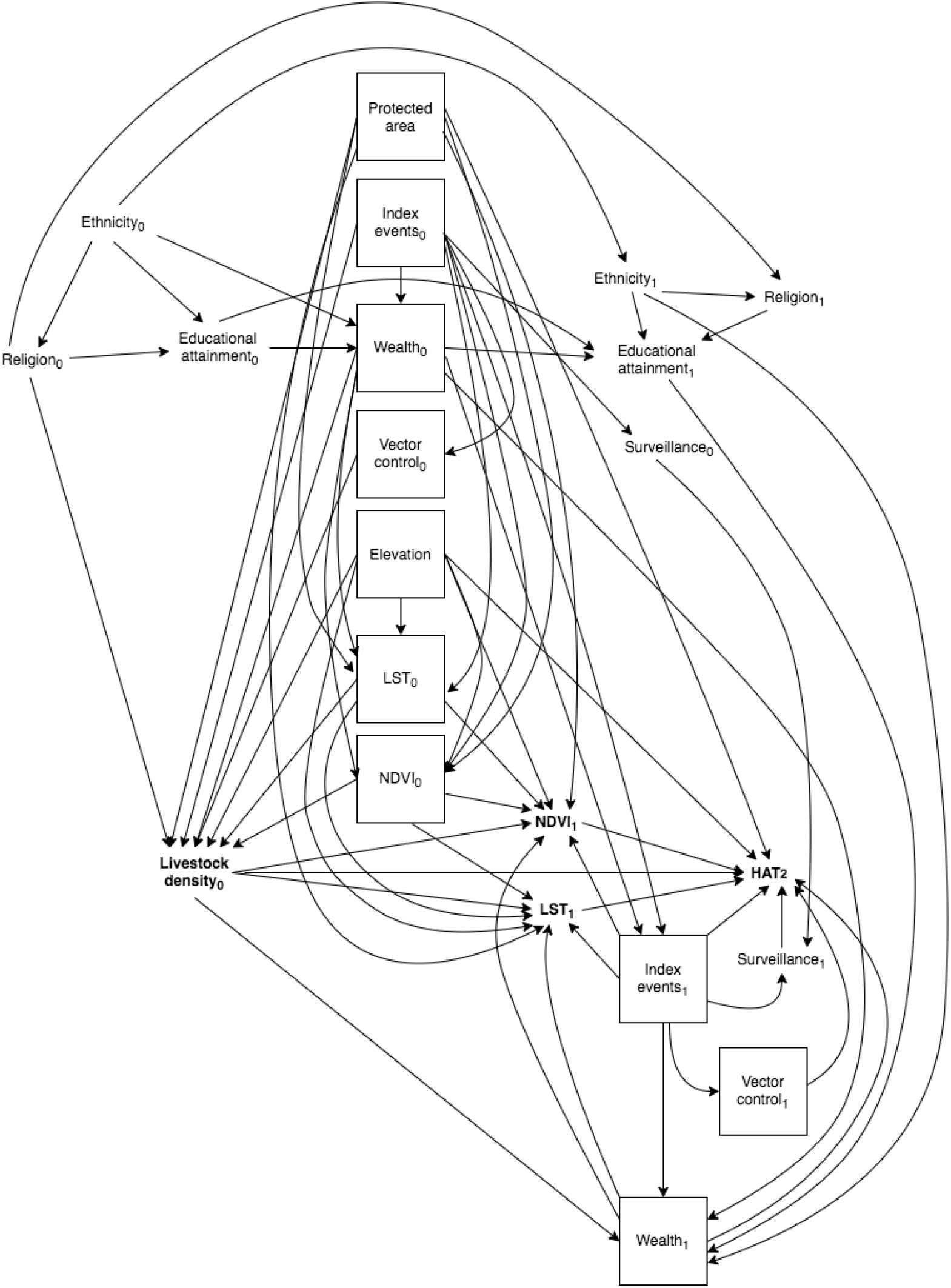
Final time-varying directed acyclic graph (DAG). Protected areas pertains to rHAT models only. Exposure and outcome of interest are bolded, and confounders in the minimally-sufficient set are denoted by solid boxes

For all time-varying variables, we assume all effects are completely mediated by the same variable’s value at the subsequent time point; for instance, the effect of index events at time 0 on HAT risk at time 2 is fully mediated by index events at time 1, with no direct effect. The exception to this is vector control, in which case we assume there is neither a direct effect on HAT at greater than one lag (i.e., no directed edge from vector control_0_ to HAT_2_), nor an indirect effect mediated by vector control at the subsequent time (i.e., no directed edge from vector control_0_ to vector control_1_).

### Measures

#### Exposure

Exposure is defined as livestock density, parameterized using a time-series of maps we have created and detailed in a separate publication linked in S1 File. We use the term “livestock” to refer collectively to cattle and pigs, however all analyses are conducted separately for each species. Livestock density is defined as the ratio of animals to humans.

For Malawi, Uganda, and DRC we generated continuous maps by year for the study period. We validated these maps using leave one out cross validation, and by comparing our final 2010 maps against the Gridded Livestock of the World 3 maps [24, 25], using WorldPop data to estimate human population and therefore density [26]. For South Sudan, given limited availability of data that provided both livestock and location data, it was only possible to produce areal (county-level) maps, and only for 2008.

#### Denominator data

Denominator data came from the University of Southhampton’s WorldPop project, which produces high resolution (1km at the equator) data on human population distributions using a flexible random forest estimation technique [27].

#### Outcome data

Outcome data included all new cases of HAT diagnosed in a given year and given cluster (Malawi, Uganda, DRC) or county (South Sudan) in the WHO Atlas of HAT [28]. Cclusters which were not represented in the Atlas and were at least 1km from any clusters with a reported case were assigned 0 cases.

#### Confounders

We estimated wealth using an exploratory factor analysis approach modeled after the DHS Wealth Index [29] but excluding livestock-related measures. We then used spatial modeling to generate continuous annual maps (detailed in S1 Appendix).

We used the EM-DAT database to identify natural disasters during the study period, and UCDP/PRIO Armed Conflict Database to identify armed conflicts [30]. Disaster was parameterized as a binary variable for our models, taking value=1 if an event occurred in a given cluster-year or county. Data on NDVI came from the Land Long Term Data Record 5 (LTDR5) [31]. Data on LST came from MODIS/Terra MOD21 for 2000-2002 [32], and from MODIS/Aqua MYD21 for 2003-2014 [33]. Elevation data came from GMTED2010 [34]; for our analyses we used the 7.5-arc-second data, which has an root mean squared error of 26-30 meters, and median elevation.

We did not attempt to adjust for vector control as top-down vector control efforts remain limited [35, 36] and it is not practical to parameterize farmer-led efforts. We do, however, present E-values on the relative risk scale to indicate the strength of uncontrolled confounding that would be required to fully explain the detected effects [37, 38].

### Parametric g-formula

In Uganda, DRC, and Malawi, we used the parametric g-formula to estimate the total effect of livestock density on HAT risk in the presence of exposure-confounder feedback. Under the potential outcomes framework for causal inference, causal estimands are defined in terms of counterfactual outcomes *Y* ^*a*^—specifically, contrasts of these counterfactual outcomes under different exposure values *Y* ^*a*^− *Y* ^*a*∗^—where *Y* denotes outcome and *A* = *a* denotes exposure fixed at level *a*. The g-formula applies standardization to estimate counterfactual outcomes, and the parametric g-formula uses parametric models to extend the g-formula to settings with numerous or high-dimensional confounders [39].

While individual-level counterfactual outcomes cannot in general be estimated from observed data, population-average counterfactual outcomes *E*[*Y* ^*a*^] can be estimated if three conditions are met: (conditional) exchangeability, positivity, and stable unit treatment value assumption (SUTVA). The identifiability criteria and implementation of the parametric g-formula are detailed in S2 Appendix. Broadly, models are fit for each time-varying confounder and for outcome at each discrete time point, and predictions generated from each model under exposure value *A* = *a*. We defined *A* = *a*\* as mean livestock density, and *A* = *a* as mean × 1.5 and specify our causal estimand on the ratio scale (S2 Appendix), generating a rate ratio analog corresponding to a 50% increase in exposure.

As the no interference assumption is violated in our study, we instead turn to partial interference, in which each unit of observation has a specified interference set *χ*_*i*_ = {*i*_1_, *i*_2_, …} [40, 41]. Partial interference is satisfied if each unit’s potential outcome is independent of the rest conditional on this interference set. Previous authors have defined the interference set in terms of its member’s counterfactual outcomes, however applications to correlation in space have been limited to linear space [41]. In our application, interference arises through movement of tsetse flies, livestock, and people. As grazing radius can vary from less than one to 10 or more kilometers per day, and tsetse flies have a range with a radius of approximately 300 meters per day, we define the interference set as all other clusters within a 5km radius [42, 43].

Validation of the parametric g-formula is conducted through implementation of the “natural course” model, whereby exposure (here, livestock density) is not fixed, but rather modeled as for time-varying confounders. The simulated outcome distribution is then compared against the observed to determine how successfully the observed distribution was recovered.

### Regression

As we did not have longitudinal data in South Sudan, exposure-confounder feedback was not a concern, allowing us to use spatial regression models rather than the parametric g-formula. Specifically, we fit Bayesian hierarchical models with ICAR and iid random effects in space (county), implemented using the R-INLA package [44].

We first fit naive Poisson models which did not account for measurement error in livestock density or wealth—that is, did not acknowledge that these variables were estimated:

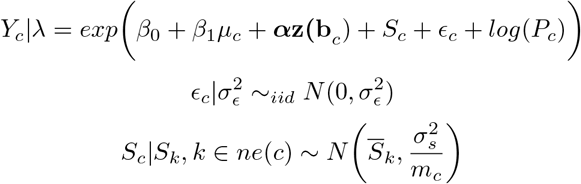

where

- *Y*_*c*_ is the number of cases in county *c*
- *µ*_*c*_ is estimated livestock (cattle or pig) density in county *c*
- ***α*** is a vector of coefficients
- **z(b**_*c*_) is a vector of confounders
- *S*_*c*_ are the structured (ICAR) spatial (county) random effects
- *ϵ*_*c*_ are the unstructured (iid) spatial (county) random effects
- *P*_*c*_ is the offset, given as population in county *c*
- *ϵ*_*c*_ are county-level iid (unstructured) random effects with variance 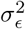
- *S*_*c*_ are county-level structured random effects which follow the ICAR model with marginal variance 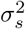
- *ne*(*c*) denotes neighbors (shared boundary) of county *c*
- *m*_*c*_ is the number of neighbors of county *c*

and ICAR is the intrinsic conditional autoregressive model, which smooths each county’s random effect to that of its neighbors, with more smoothing performed for counties with fewer neighbors. We parameterized exposure (livestock density) such that exponentiated effect estimates could be interpreted as the rate ratio for a 50% increase in density, allowing direct comparison with g-formula results in the other countries. We used penalized complexity (PC) priors for the smoothing model, with *Pr*(*σ*_*ϵ*_) *>* 1 = *Pr*(*σ*_*s*_) *>* 1 = 0.01. This yields a posterior 99% credible interval for each random effect’s residual rate ratio of (0.36, 2.71) [45, 46].

Next, we re-fit these models, this time specifying livestock density and wealth as random effects following a classical measurement error (MEC) model (implementing these models within the g-formula was not practical for the other study countries). Under this model, the true value *x* cannot be observed directly, but is instead measured with error. We detail implementation of this model in S3 Appendix.

As a sensitivity analysis, we also repeated these analyses restricted to the active surveillance data, as we expect bias in HAT reporting to follow different mechanisms in the active surveillance versus full dataset.

### Data and code availability

HAT outcome data can be requested from the WHO (https://www.who.int/trypanosomiasis_african/country/foci_AFRO/en/) and livestock density maps can be downloaded from https://github.com/JulianneMeisnerUW/LivestockMaps. Confounder data are available for download from the websites linked in our References. All analyses were performed in R, and all code are available in the same GitHub repository.

## Results

### Descriptive statistics

After removing predicted clusters within within 1km of locations in the WHO Atlas of HAT data and more than 5 hours’ travel time from fixed health facilities capable of HAT diagnosis, we were left with 3,982 clusters/year in Malawi, 5,746 clusters/year for gHAT models in Uganda, 13,570 clusters/year for rHAT models in Uganda, and 247,205 clusters/year in DRC. In South Sudan, the study area was comprised of 17 counties, with active surveillance being performed in 14 of them in 2008. General study areas are presented in Fig 4. Note this figure represents the administrative areas represented in the study data in each country, however in Malawi, Uganda, and DRC, the study area is defined at the cluster-level, not the areal-level (i.e., not all clusters within a given administrative area highlighted in Fig 4 appear in the study data).

**Fig 4.**
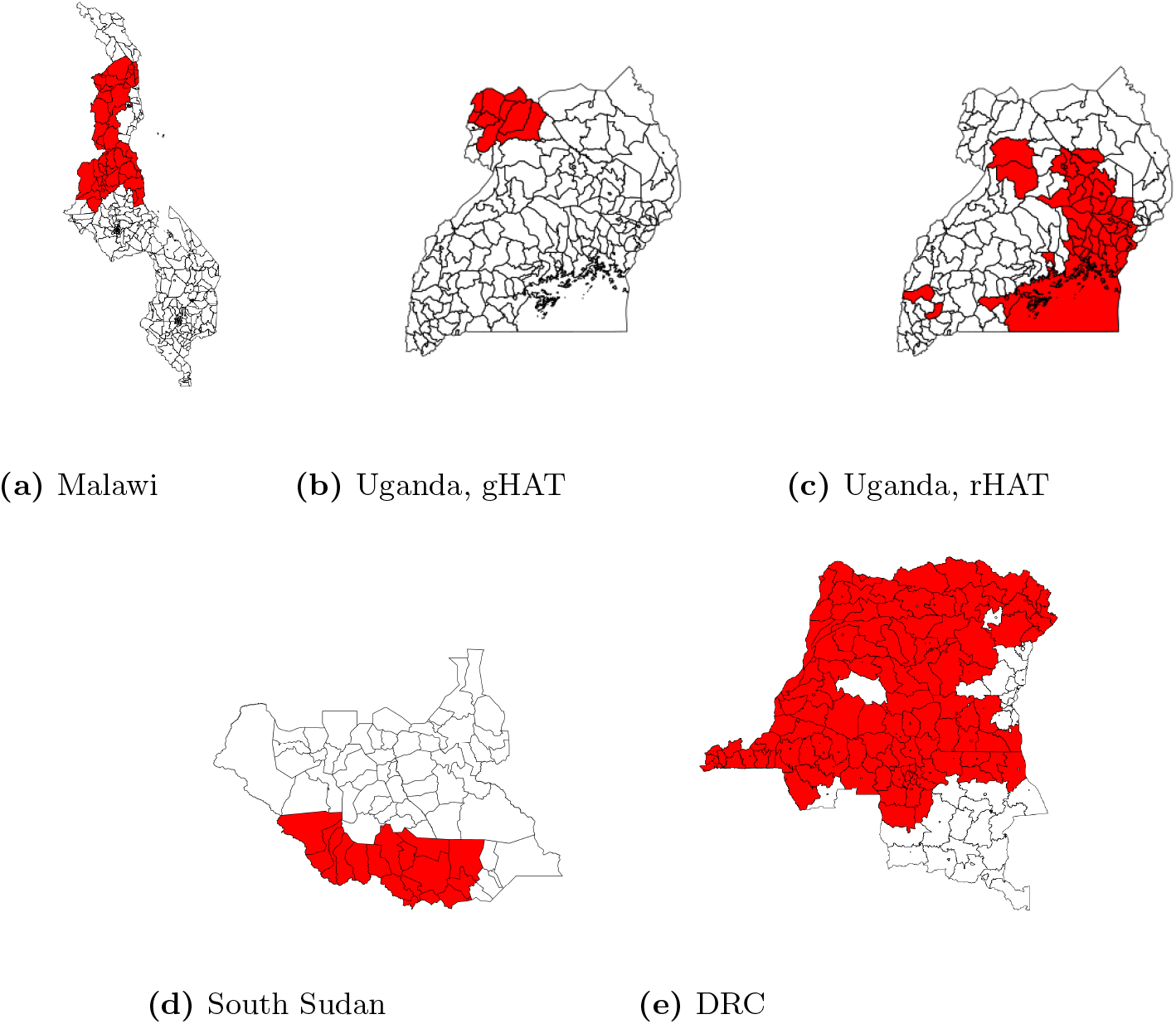
National maps with administrative areas represented in the study data highlighted in red. (a) Malawi: traditional authority (administrative level 3); (b-d) Uganda and South Sudan: county (administrative level 2); (e) DRC: territory (administrative level 2)

No armed conflicts in Malawi appeared in the UCDP/PRIO database during the study period, and no natural disasters in South Sudan appeared in the EM-DAT database in 2008. Descriptive statistics are presented in Tables 1 and 2, with additional descriptive statistics figures presented in S4 Appendix, including temporal trends (aggregated over clusters) and maps for key variables.

**Table 1.** Descriptive statistics, Malawi, Uganda, and DRC.

| Variable | Mean (sd) |  |  |  |
| --- | --- | --- | --- | --- |
|  | Malawi | Uganda, gHAT | Uganda, rHAT | DRC |
| Cattle density | 0.09 (0.31) | 0.31 (0.92) | 0.16 (0.42) | 0.06 (0.06) |
| Pig density | 0.09 (0.19) | 0.24 (0.72) | 0.06 (0.10) | 0.06 (0.03) |
| Elevation (meters) | 1,125 (280) | 890 (166) | 1,099 (79) | 524 (194) |
| HAT cases | 0.01 (0.14) | 0.04 (0.35) | 0.01 (0.15) | 0.02 (0.35) |
| LST (K) | 307 (4.7) | 281 (96) | 277 (95) | 306 (3.67) |
| NDVI | 0.15 (0.14) | 0.28 (0.13) | 0.26 (0.21) | 0.22 (0.18) |
| Wealth | 1.30 (1.04) | 0.84 (0.03) | 0.85 (0.03) | 0.25 (0.22) |
| Conflict | 0 (0%)* | 743 (68%)* | 1,824 (71%)* | 177,468 (9%)* |
| Disaster |  |  |  |  |
| Flood | 19,548 (33%)* | 413 (38%)* | 2,290 (0.9%)* | 46,921 (2.37%)* |
| Storm | 1,213 (2%)* | 0 (0%)* | 0 (0%)* | 0 (0%)* |
| Epidemic | 0 (0%)* | 2,065 (1.9%)* | 6,307 (2.5%)* | 28,343 (1.43%)* |
| Landslide | 0 (0%)* | 0 (0%)* | 127 (0.05%)* | 0 (0%)* |
| Drought | 0 (0%)* | 0 (0%)* | 8 (<0.01%)* | 0 (0%)* |
| Earthquake | 0 (0%)* | 0 (0%)* | 0 (0%)* | 410 (0.02%)* |
| Wildfire | 0 (0%)* | 0 (0%)* | 0 (0%)* | 16,575 (0.84%)* |

Descriptive statistics over study clusters and period (2000-2014 for Malawi, 2000-2018 for Uganda and DRC). sd: standard deviation. \*n(%)

**Table 2.** Descriptive statistics, South Sudan.

| Variable | Mean (sd) |
| --- | --- |
| Cattle density | 0.77 (1.15*) |
| Pig density | 0.02 (1.25*) |
| Elevation (meters) | 693 (158) |
| HAT cases | 34 (52) |
| Number screened | 2,953 (4,200) |
| LST (K) | 312.54 (3.22) |
| NDVI | 0.28 (0.07) |
| Wealth | 0.19 (0.004*) |
| Conflict | 8 (47%)** |

Descriptive statistics over study counties. 2008 data: cattle density, pig density, HAT cases, number screened, WorldPop population, LandScan population, wealth. 2007 data: LST, NDVI. 2006 data: conflicts. sd: standard deviation. \*Mean of design-based standard error.\*\*n, (%)

### Parametric g-formula

In Uganda, for the rHAT models the mean of the squared difference between observed HAT cases and that predicted by the “natural course” model across cluster-years was less than 1 × 10^−4^ for both cattle and pigs. For the gHAT models, the pig models performed very badly in 2010 and 2012; removing these years from all subsequent analyses for pigs, this value was *<* 0.0005 for both cattle and pigs. In Malawi and DRC, this value was *<* 1 × 10^−4^ for both species across all years.

In the Malawi models, several confounders needed to be dropped due to the very low number of observed cases (as low as 18 cases reported in 2012) and resulting concerns surrounding model over-fitting. Namely, location within a protected area, elevation, and disasters were removed from the Malawi models. LST was furthermore dropped from all analyses due to very large levels of missingness.

Results from the parametric g-formula are presented in Table 3. We found a 50% increase in cattle density was associated with a 61% higher risk of rHAT in Uganda (95% CI 0.90, 2.99) and a 12% lower risk of rHAT in Malawi (95% CI 0.71, 1.10). For gHAT, a 50% increase in cattle density was associated with and a 12% lower risk of gHAT in Uganda (95% CI 0.50, 1.77) and a 17% higher risk of gHAT in DRC (95% CI 1.04, 1.32). For pigs, a 50% increase in density was associated with a 107% higher rHAT risk in Uganda (95% CI 1.15, 2.75) and a 58% lower rHAT risk in Malawi (95% CI 0.23, 0.67). For gHAT, a 50% increase in pig density was associated with and a 102% higher risk of gHAT in Uganda (95% CI 0.87, 3.94) and a 23% higher risk of gHAT in DRC (95% CI 1.10, 1.37).

**Table 3.**
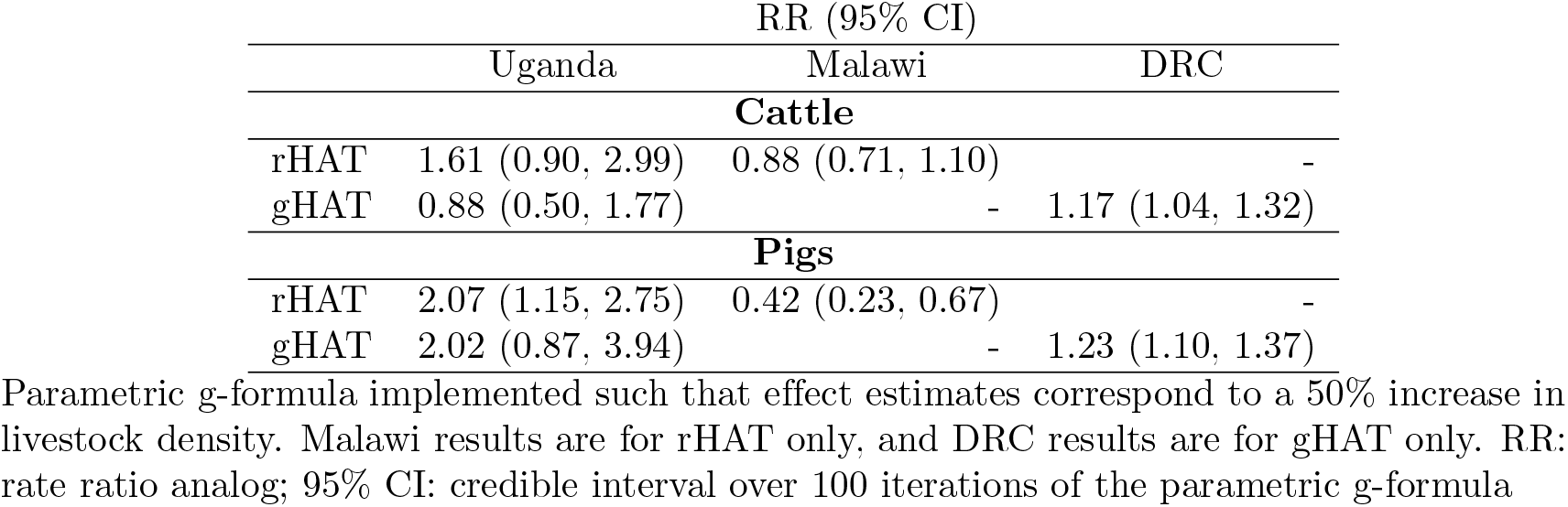
Parametric g-formula results.

| RR (95% CI) |  |  |  |
| --- | --- | --- | --- |
|  | Uganda | Malawi | DRC |
| Cattle |  |  |  |
| rHAT | 1.61 (0.90, 2.99) | 0.88 (0.71, 1.10) | - |
| gHAT | 0.88 (0.50, 1.77) | - | 1.17 (1.04, 1.32) |
| Pigs |  |  |  |
| rHAT | 2.07 (1.15, 2.75) | 0.42 (0.23, 0.67) | - |
| gHAT | 2.02 (0.87, 3.94) | - | 1.23 (1.10, 1.37) |

Parametric g-formula implemented such that effect estimates correspond to a 50% increase in livestock density. Malawi results are for rHAT only, and DRC results are for gHAT only. RR: rate ratio analog; 95% CI: credible interval over 100 iterations of the parametric g-formula

### Regression

Total effect estimates are presented in Table 4. While both results are presented, we will only discuss results from the measurement error model as the naive model estimates are biased. After adjustment for confounders {wealth, NDVI, LST, elevation, armed conflict}, the rate ratio (RR) was 0.63 (95% CI 0.54, 0.77) for cattle, and 0.66 (0.50, 0.98) for pigs. Cattle effect estimates were slightly attenuated and no longer significant when restricted to active surveillance data (RR 0.79, 95% CI 0.35, 1.20), while pig estimates were not appreciably changed (RR 0.59, 95% CI 0.36, 0.91).

**Table 4.** Total effect results, South Sudan.

| Denominator | Model | RR (95% CI) |
| --- | --- | --- |
| **Cattle** |  |  |
| WorldPop | Naive | 0.59 (0.37, 0.83) |
| WorldPop | MEC | 0.63 (0.54, 0.77) |
| Number sampled | Active surveillance | 0.79 (0.35, 1.20) |
| **Pigs** |  |  |
| WorldPop | Naive | 0.63 (0.31, 1.18) |
| WorldPop | MEC | 0.66 (0.50, 0.98) |
| Number sampled | Active surveillance | 0.59 (0.36, 0.91) |

Posterior mean rate ratios and 95% credible intervals for livestock density in South Sudan, after adjustment for wealth, NDVI (lagged 1 year), LST (lagged 1 year), elevation, and armed conflict. Density is parameterized such that rate ratios correspond to a 50% increase in density. RR: rate ratio; CI: credible interval; Naive: models which do not account for measurement error in wealth or livestock density; MEC: measurement error models

### E-values

E-values are presented in Table 5. In all cases, strong unmeasured confounding would be required to explain our findings. This is particularly true for the cattle-rHAT association in Uganda, the pig-rHAT association in Uganda and Malawi, the pig-gHAT association in Uganda and South Sudan, and the cattle-gHAT association in South Sudan.

**Table 5.** E-value results.

|  | Uganda | Malawi | DRC | South Sudan |
| --- | --- | --- | --- | --- |
| **Cattle** |  |  |  |  |
| rHAT | 2.6 | 1.53 | - | - |
| gHAT | 1.53 | - | 1.62 | 2.55 |
| **Pigs** |  |  |  |  |
| rHAT | 3.56 | 4.19 | - | - |
| gHAT | 3.46 | - | 1.76 | 2.4 |

The strength, on the relative risk or rate ratio scale, of the relationship between an unmeasured confounder and exposure (livestock density) and between an unmeasured confounder and outcome (HAT risk) that would be required to fully explain the estimated effect. South Sudan results are presented for measurement error models.

### Simulations

While the finding of a protective effect in Malawi for both cattle and pigs has several possible explanations other than bias—detailed below in our Discussion section—bias due to failure of passive surveillance systems resulting in underreporting is a significant concern. Similarly, the finding of a possible reservoir effect of cattle for gHAT in DRC is surprising and inconsistent with the gHAT findings in Uganda and South Sudan, and may be explained by underreporting due to failure of passive surveillance systems, or to undercoverage of active surveillance systems in high-risk foci in conflict zones or remote locations.

We thus conducted a simulation study to explore the degree of underreporting that would be needed to “explain away” these findings. For DRC, all scenarios followed a model in which a 50% increase in cattle density causes a 20% decrease in gHAT risk, and the same increase in pig density causes a 15% decrease in gHAT risk. These scenarios differed by the level of underreporting: one unreported case per 10 reported cases, per three reported cases, and per two reported cases. In Malawi, unreported cases followed a model in which a 50% increase in cattle density causes a 20% increase in rHAT risk, and the same increase in pig density causes a 15% increase in risk; underreporting levels were simulated at one half, three, and 10 unreported cases per one reported case. We simulated a relatively higher underreporting level in Malawi (i.e., a max of 10 unreported cases per reported cases versus a max of 0.5 unreported cases per reported case) due to the absence of active surveillance activities in Malawi and evidence that underreporting is much higher for rHAT than gHAT foci.

Results are presented in Table 6. We found that under the modeled scenarios, a relatively high level of underreporting would be required to explain our results in these two countries. In Malawi, three unreported cases per one reported case would be required to result in a protective effect when indeed cattle play a reservoir role among the unreported cases; for pigs, an even higher level of underreporting would be required. In DRC, negative (protective) effects were not recovered under any of the simulated levels of underreporting. Note that in Malawi the highest level of underreporting corresponds to 4,122 unreported cases, while in DRC this level corresponds to 18,135 unreported cases.

**Table 6.** Simulation results.

| Underreporting ratio | Cattle | Pigs |
| --- | --- | --- |
| Malawi |  |  |
| 0.5 | 0.94 (0.77, 1.09) | 0.55 (0.36, 0.87) |
| 3 | 1.03 (0.91, 1.16) | 0.92 (0.62, 1.36) |
| 10 | 1.19 (1.10, 1.32) | 1.49 (1.06, 2.05) |
| DRC |  |  |
| 0.1 | 1.14 (1.03, 1.26) | 1.01 (0.99, 1.06) |
| 0.3 | 1.17 (1.07, 1.25) | 1.03 (1.00, 1.11) |
| 0.5 | 1.17 (1.08, 1.27) | 1.04 (1.00, 1.11) |

Results from implementation of the parametric g-formula under a variety of simulated underreporting scenarios, each implemented such that effect estimates correspond to a 50% increase in livestock density. Underreporting ratio calculated as # unreported cases / # reported cases. Malawi results are for rHAT only, and DRC results are for gHAT only. RR: rate ratio analog; 95% CI: credible interval over 100 iterations of the parametric g-formula.

## Discussion

In Uganda we found cattle and pigs both have a positive effect on rHAT risk. For gHAT, we found cattle have negative (protective) effect, while pigs have a positive (harmful) effect. In DRC, we detected a positive effect on gHAT for both cattle and pigs, being marginally stronger for pigs than for cattle. Results did reach statistical significance in Uganda.

In Malawi (rHAT) and South Sudan (gHAT), we found both cattle and pigs have protective (negative) effects, with this result being stronger in pigs than in cattle in Malawi and cattle results not reaching statistical significance in this country. In South Sudan, restricting analyses to the active surveillance data did not appreciably change our results.

These results indicate the zooprophylactic effect—whereby livestock exert a protective effect on HAT risk due to tsetse preference for animal hosts over humans—outweighs the reservoir effect for both cattle and pigs in Malawi and South Sudan and for cattle in gHAT foci in Uganda, however the reservoir effect outweighs the zooprophylactic effect for pigs in gHAT foci in both DRC and Uganda, for cattle in gHAT foci in DRC, and for pigs and cattle in rHAT foci in Uganda. Comparison of results across cattle versus pigs should also consider our choice of a relative parameterization for exposure; that is, a 50% increase in pig density is not equivalent, in absolute terms, to a 50% increase in cattle density, within and across foci.

Our findings initially appear discordant with mechanistic knowledge about HAT transmission, i.e., cattle and pigs are reservoirs for rHAT, and as the gHAT focus in Uganda is contiguous with that in South Sudan, transmission dynamics should be nearly equivalent. However, it is important to remember these results reflect complex epidemiological factors that extend beyond transmission mechanisms to local population dynamics and control measures. For instance our Malawi results do not conflict with a scenario in which cattle and pigs are theoretical reservoirs of rHAT, however the wildlife reservoir so markedly dominates these domestic animal reservoirs that the zooprophylactic effect prevails. This could arise if the predominant vector species (*Glossina morsitans morsitans*) exhibits a strong preference for wildlife, or if domestic animal reservoirs are adequately controlled through use of insecticides or trypanocides, or environmental measures such as brush clearing. Similarly, differences in host abundance and control measures between Uganda and South Sudan could explain the discrepancies between the results observed in these two countries. This is particularly relevant for the case of pigs; as South Sudan is a predominantly Muslim country, very few pigs are kept, and the sociocultural characteristics of higher pig density clusters are likely highly divergent from those in low pig density clusters, potentially driving similar divergence in HAT epidemiology. Presence of a zooprophylactic effect in South Sudan and for cattle in gHAT foci in Uganda is consistent with evidence from southeastern Uganda that *Glossina fuscipes fuscipes*, the predominant gHAT vector in South Sudan and Uganda, exhibits strong zoophilic behavior [47]. Note that while there are several possible mechanisms which could give rise to this biting preference (i.e., relative abundance of host species, preferences inherent to fly species, etc.), pointing to different opportunities for intervention, all are expected to result in a net negative effect of livestock density on HAT risk. The finding of a positive cattle-gHAT effect in DRC but not Uganda or South Sudan is surprising, and of uncertain significance.

However, our results could, of course, be subject to bias. With regards to confounding, in Uganda, DRC, and South Sudan we feel we have achieved reasonable control for all confounders except vector control efforts. We would expect vector control to have a positive effect on livestock density (as AAT control would increase livestock density) and a negative effect on HAT risk (through decreased cases), thus unmeasured confounding by vector control would bias effect estimates towards zero, making it harder to detect positive (*>* 1) effects, and easier to detect negative (*<* 1) effects. Thus, it is possible failure to adjust for vector control explains our the negative effects we are attributing to zooprophylaxis, however the zooprophylactic effect was most pronounced in foci with limited top-down vector control efforts (Malawi and South Sudan). This, along with our calculated E-values, indicates unmeasured confounding by vector control may explain the rHAT effect for cattle in Malawi and the gHAT effect for cattle in Uganda, but is unlikely to be responsible for the other effect estimates. Furthermore, we approximated wealth, a latent construct, using factor analysis (detailed in S1 Appendix), which may not capture the dimension of wealth which is most relevant to our study, driving residual confounding by this variable. Finally, in Malawi we did not adjust for several known confounders due to overfitting concerns. Again, our E-values indicate this unmeasured confounding may explain the cattle results in Malawi, but are unlikely to explain the pig results.

A second major source of bias is selection bias. In Malawi, Uganda, and DRC, study area was defined by proximity to a fixed health facility capable of HAT diagnosis, however we did not have longitudinal data for this variable. In South Sudan, we defined study counties by appearance in the WHO Atlas of HAT dataset, and thus underreporting of the outcome (discussed below) may drive selection bias. In both cases, selection bias arises if excluded clusters or counties differ systematically in their distribution of livestock density or confounders from the included counties or clusters. While we attempted to evaluate the underreporting as the driver of our more surprising findings in Malawi and DRC via simulation studies, the simulated scenarios were somewhat simple, governing only the mechanism by which unreported cases were generated.

As with all research, measurement error threatens the validity of these findings. We attempted to account for measurement error arising from estimation of wealth and livestock density in South Sudan, however these measurement error models do not account for errors in the raw data used for livestock or wealth mapping, for instance a systematic under-reporting of livestock ownership due to concerns about increased taxation. Furthermore, we did not implement measurement error models in the remaining study countries, however we did restrict analyses to years bounded by those with available input data used to generate livestock density estimates (see S1 File for detail). We expect such measurement error to be non-differential with respect to HAT risk, biasing our effect estimates towards the null in expectation. With regards to LST, NDVI, index events, and elevation, measurement error either in the attribute itself, or in its geolocation, will drive residual confounding and bias our effect estimates.

We also expect measurement error in the outcome. As with all surveillance data, reporting of HAT cases is likely to be incomplete, and some HAT foci have no effective surveillance systems [10]. This is particularly true for data collected via passive surveillance, which accounts for all of the Malawi data and Uganda rHAT data. Under-reporting in the passive surveillance data arises from cases who do not seek care, seek care but are not correctly diagnosed, or are diagnosed but not reported [48]. Under-reporting in active surveillance data arises from lack of surveillance in remote or politically unstable foci [49], and incomplete coverage of populations in which active surveillance occurs [50]. If under-reporting is random with regards to space and either HAT risk or livestock density, power will be compromised but effect estimates will be unbiased in expectation. As we expect comparatively less underreporting in the active surveillance data than the passive surveillance data, the fact that our findings in South Sudan were not markedly different when restricted to active surveillance cases alone is reassuring.

Ecological bias may also compromise the validity of our findings. Cross-level (ecological) bias may result from (1) an unmeasured variable which is either (a) an individual-level risk factor whose association with exposure differs across groups, or (b) an individual-level effect modifier whose distribution or effect differs across groups [51]; or (2) aggregation of variables (e.g., from the household to the cluster) [52, 53], termed “pure specification bias” [54]. While wealth was the only confounder measured at the household-level, elevation, NDVI and LST were aggregated from the resolution at which they were defined in the source data to our unit of analysis. Furthermore, analysis of contextual (inherently group-level) variables is not immune to ecological bias [55]. As the size of the ecological association is sensitive to the level of aggregation (being attenuated at higher levels of aggregation), we expect our South Sudan results to be more sensitive to this source of bias than those from Malawi, Uganda, and DRC [56]. Furthermore, findings from ecological studies are sensitive to the grouping definition used [55]; in Malawi, Uganda, and DRC grouping was uniform across space (defined by grid cells), however in South Sudan we utilized administrative boundaries to define groups, which are uneven and do not have meaning to our research question.

Finally, our results are subject to the g-null paradox. When the parametric g-formula is implemented using non-saturated models and the vector of confounders has any discrete components, as is the case in our study, not all models can be correctly specified. While this may result in the null hypothesis being falsely rejected in theory, in practice the g-formula has been able to estimate null effects, indicating the bias induced by the g-null paradox is relatively small compared with random variance [57].

## Conclusion

Our greatest concerns for the validity of these findings are residual confounding by vector control, overfitting of the Malawi models, ecological bias in the South Sudan results, and underreporting in the outcome data. Despite these weaknesses, our study has taken a novel approach to studying the impact of livestock density on HAT risk in a real-world context in which transmission mechanisms exert their effects in the face of spatially-dynamic intervention deployment and vector ecology and behavior, indicating domestic animals are important reservoirs of rHAT in Uganda but not Malawi, and pigs—and possibly cattle—may be reservoirs for gHAT. While achieving 2020 targets for EPHP had previously appeared within reach, lapses in control activities due to COVID-19, and long-standing concerns regarding persistent foci and cryptic animal reservoirs, render the need to close HAT knowledge gaps addressed by this research as pressing as ever.

## Data Availability

All relevant data are within the manuscript and its Supporting Information files.

## Supporting information

**S1 Appendix. Wealth mapping**. Methods and results

**S2 Appendix. Motivation and implementation of the parametric g-formula**.

**S3 Appendix. Implementation of the measurement error model in South Sudan**.

**S4 Appendix. Descriptive statistics figures**.

**S1 File. Livestock mapping manuscript**. PDF of author manuscript containing detail on methodology for livestock mapping.

## Acknowledgments

The authors wish to express sincere appreciation to the World Health Organization’s Department of Control of Neglected Tropical Diseases for use of their Atlas of HAT data, and to the HAT research community, in particular Prof. Eric Fevré and Dr. Richard Selby.

## S1 Appendix: Wealth mapping

### Data collection and processing

Data sources used for livestock and wealth mapping are presented in Table 1.1. There were 10,330 clusters in the final dataset for Malawi, 4,361 in Uganda, and 1,164 in DRC. After reading in data, we harmonized variables so that the wealth index could be calculated across surveys (as detailed in the following section), allowing for smoothing in time.

**Table 1.1:**
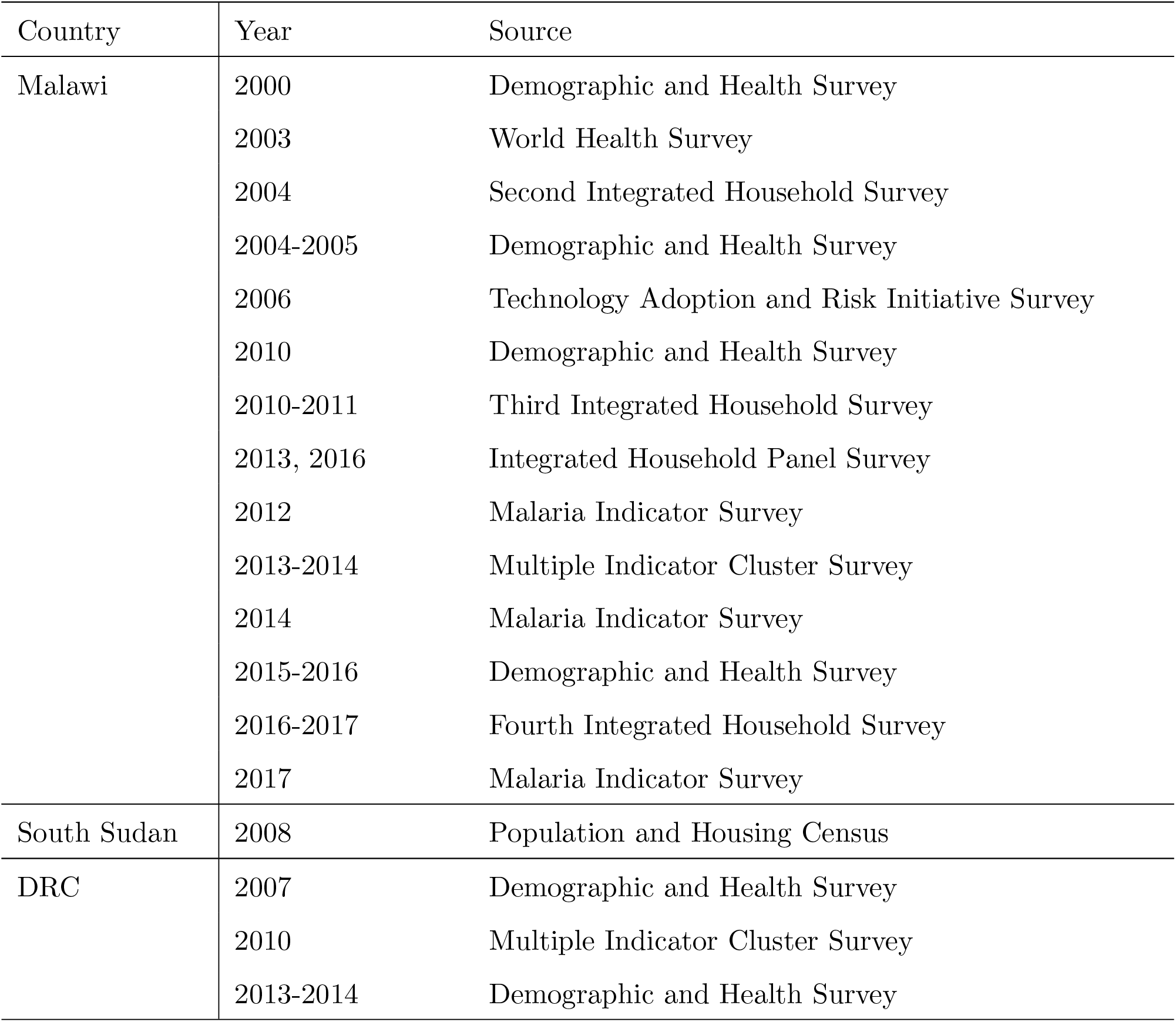
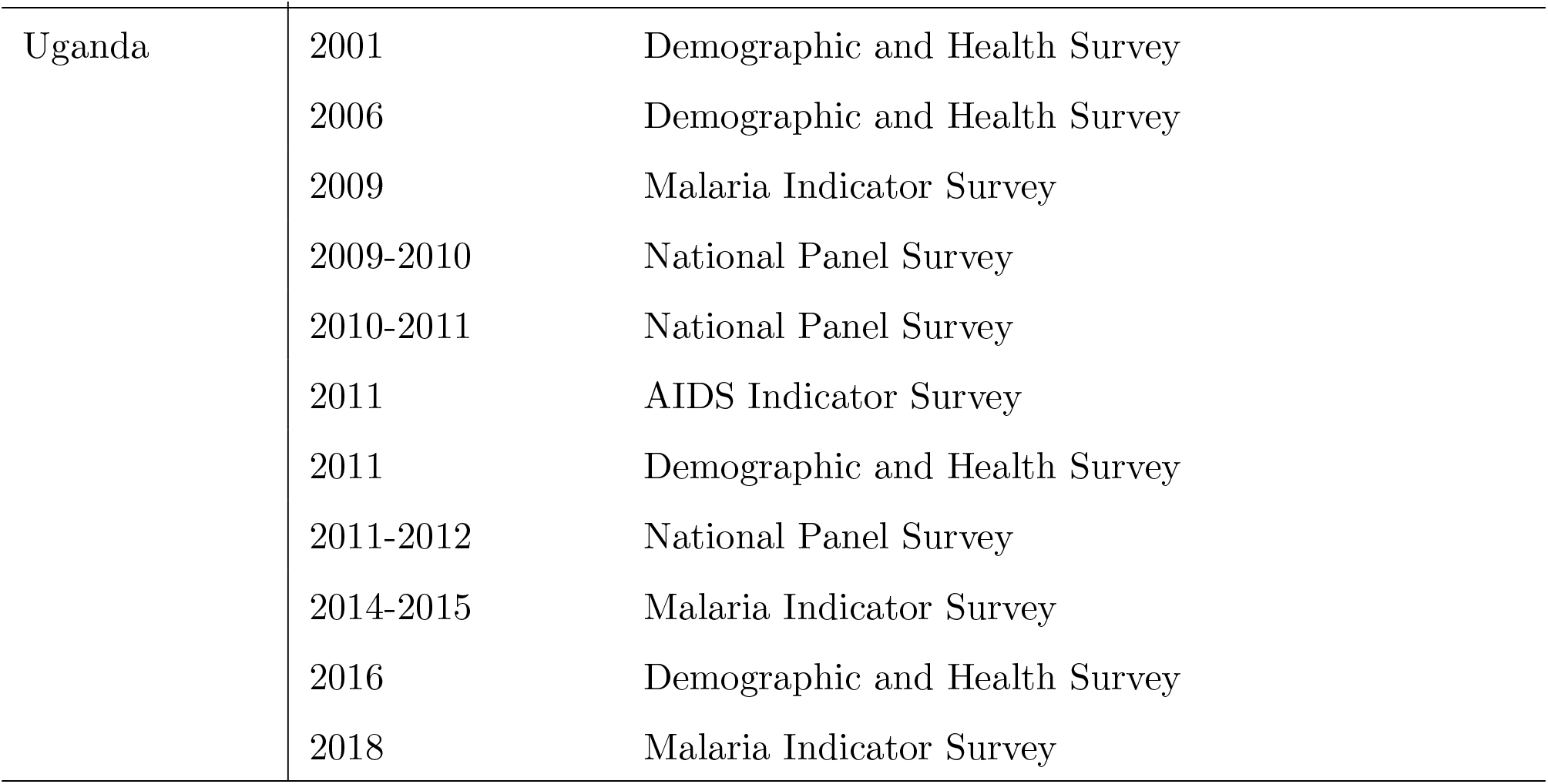
Data sources by country

| Country | Year | Source |
| --- | --- | --- |
| Malawi | 2000 | Demographic and Health Survey |
|  | 2003 | World Health Survey |
|  | 2004 | Second Integrated Household Survey |
|  | 2004-2005 | Demographic and Health Survey |
|  | 2006 | Technology Adoption and Risk Initiative Survey |
|  | 2010 | Demographic and Health Survey |
|  | 2010-2011 | Third Integrated Household Survey |
|  | 2013, 2016 | Integrated Household Panel Survey |
|  | 2012 | Malaria Indicator Survey |
|  | 2013-2014 | Multiple Indicator Cluster Survey |
|  | 2014 | Malaria Indicator Survey |
|  | 2015-2016 | Demographic and Health Survey |
|  | 2016-2017 | Fourth Integrated Household Survey |
|  | 2017 | Malaria Indicator Survey |
| South Sudan | 2008 | Population and Housing Census |
| DRC | 2007 | Demographic and Health Survey |
|  | 2010 | Multiple Indicator Cluster Survey |
|  | 2013-2014 | Demographic and Health Survey |
| Uganda | 2001 | Demographic and Health Survey |
|  | 2006 | Demographic and Health Survey |
|  | 2009 | Malaria Indicator Survey |
|  | 2009-2010 | National Panel Survey |
|  | 2010-2011 | National Panel Survey |
|  | 2011 | AIDS Indicator Survey |
|  | 2011 | Demographic and Health Survey |
|  | 2011-2012 | National Panel Survey |
|  | 2014-2015 | Malaria Indicator Survey |
|  | 2016 | Demographic and Health Survey |
|  | 2018 | Malaria Indicator Survey |

### Generating the wealth index

Our process for wealth mapping followed that laid out in the DHS Wealth Index [1] handbook, with the exception of exclusion of livestock-related variables in our wealth index. The final variables included in our wealth models were, defined at the level of the household (M: Malawi, U: Uganda, S: South Sudan, D: DRC):

- Presence of a domestic servant *(M, U, D)*
- Ownership of agricultural land *(M, U, S, D)*
- Amount of land owned (converted to acres) *(M, U, S, D)*
- Ownership of dwelling *(M, U, S, D)*
- Type of fuel used for cooking *(M, U, S, D)*
- Water source *(M, U, S, D)*
- Toilet type *(M, U, S, D)*
- Whether toilet is shared with other households *(M, U, D)*
- Main floor material of dwelling *(M, U, D)*
- Main material of dwelling walls *(M, U, D)*
- Roof material *(M, U, D)*
- Presence of electricity *(M, U, D)*
- One or more household members has a bank account*(M, U, D)*
- Vehicle (none; animal cart or bicycle; motorcycle or scooter; car, mini-bus or truck/lorry) *(M, U, D)*
- Number of rooms per household member *(M, U, S, D)*
- Household owns one or more:
  - Chairs *(M, U, D)*
  - Table *(M, U)*
  - Clock or watch *(M, U, D)*
  - Bucket *(M)*
  - Clothes washing machine *(M)*
  - Dish washing machine *(M, U, D)*
  - Refrigerator *(M, U, S, D)*
  - Mobile phone *(M, U, S, D)*
  - Landline (non-mobile phone) *(M, U, S, D)*
  - Television *(M, U, S, D)*
  - Computer *(M, U, S, D)*
  - Radio *(M, U, S, D)*
  - Sewing machine *(M, D)*
  - Bed *(M, U, D)*
  - Upholstered couch, sofa, chairs, or set *(M, U)*
  - Paraffin or kerosene lamp *(M, U, D)*
  - Boat *(M, U, S, D)*
  - Fishing net *(M)*
  - Mortar and pestle *(M)*
  - Fan *(M, S*
  - Air conditioning *(M)*
  - Machine to play cassette tapes, CDs, DVDs, or Hi-Fi *(M, U)*
  - VCR *(Ms)*
  - Kerosene or paraffin stove *(M)*
  - Electric or gas stove/hot plate *(M, D)*
  - Beer-brewing drum *(M)*
  - Coffee table for sittingroom *(M)*
  - Cupboards, drawers, bureau *(M, U)*
  - Desk *(M)*
  - Iron for pressing clothes *(M)*
  - Satellite dish *(M, S)*
  - Solar panel *(M, U)*
  - Generator *(M, U, D)*
  - Plough *(M)*
  - Tractor *(S)*
  - Axe or hoe *(D)*
- Proportion of possessions enumerated by the survey in question that the household owns *(M, U, D)*

We coded all variables such that a lower level corresponded to lower wealth, and a higher lever corresponded to greater wealth (e.g., we used the variable number of rooms per household member rather than number of household members per room, as the latter would have a negative association with wealth). We assigned levels to categorical variables as detailed in Tables 1.2-1.7; note apparent repetition within variable levels reflect different categorizations across surveys.

### Factor analysis

For Malawi, Uganda, and DRC, we performed exploratory factor analysis separately for each of three periods, to reflect the expectation that factor loadings would change over time (e.g., in early periods ownership of a mobile phone would be much more strongly associated with wealth than in later periods): 2000-2006, 2006-2012, and 2012-2020. For South Sudan, data were only available at the county-level and for a single year (2008), thus wealth mapping was conducted at the county-level for 2008.

First, we removed all variables with more than 5% missingness, and all variables with standard deviation equal to 0. We then created a correlation matrix using the mixedCor function in the psych package in R [2], and performed exploratory factor analysis using the fa() function in the same package, setting our arguments to those specified by the DHS Wealth Index manual: principal components extraction with one factor extracted, imputation of mean for missing data, and estimation of the factor scores using the regression method.

**Table 1.2:** Levels for cooking fuel

| Level | Fuel types |
| --- | --- |
| Malawi, Uganda., DRC |  |
| 4 | Electricity |
| 3 | Gas, kerosene, liquefied petroleum gas, biogas, natural gas, paraffin, oil, solar |
| 2 | Charcoal, coal, lignite |
| 1 | Wood, firewood (collected or purchased) |
| 0 | Dung, straw, shrub, grass, crop residue, saw dust |
| South Sudan |  |
| 4 | Electricity |
| 3 | Gas |
| 2 | Charcoal |
| 1 | Firewood |
| 0 | Cow dung, Grass |

**Table 1.3:** Levels for water source

| Malawi, Uganda, DRC |  |
| --- | --- |
| Level | Water source |
| 4 | Piped into own dwelling, compound, yard or plot |
| 3 | Public or communal standpipe/tap, piped into neighbor's dwelling |
| 2 | Private (in own dwelling, yard or plot) or public protected well, tube well, borehole, protected spring, rain tank or rainwater, channeled by gravity flow scheme |
| 1 | Tanker truck/bowser, cart with small tank, bottled or sachet water, water vendor |
| 0 | Unprotected well or spring, private (in own dwelling, yard or plot) or public open well, river, dam, lake, pond, stream, canal, irrigation channel |
| South Sudan |  |
| 3 | Water filtering stations with common standpipe, mechanical boreholes with common standpipe, sand filter with common standpipe, |
| 2 | Deep borehole with network, deep borehole without network, hand pump, water vendor from deep borehole |
| 1 | Shallow well, hafeer/dam with filter, water vendor from shallow well/pond/river/spring |
| 0 | Hafeer/dam without filter, still (turda/fula/river) or running (river/pond/tura'a) open water source |

**Table 1.4:** Levels for toilet type

| Level | Toilet type |
| --- | --- |
| Malawi, Uganda, DRC |  |
| 3 | Flush toilet (piped, pour flush, or unspecified) |
| 2 | VIP latrine, composting toilet, Ecosan toilet, latrine with slab and/or roof |
| 1 | Latrine without slab or roof, public pit toilet or latrine, bucket, hanging toilet |
| 0 | No facilities/open, bush, field |
| South Sudan |  |
| 5 | Private flush toilet |
| 4 | Shared flush toilet |
| 3 | Private pit latrine |
| 2 | Shared pit latrine |
| 1 | Bucket |
| 0 | No facilities |

**Table 1.5:**
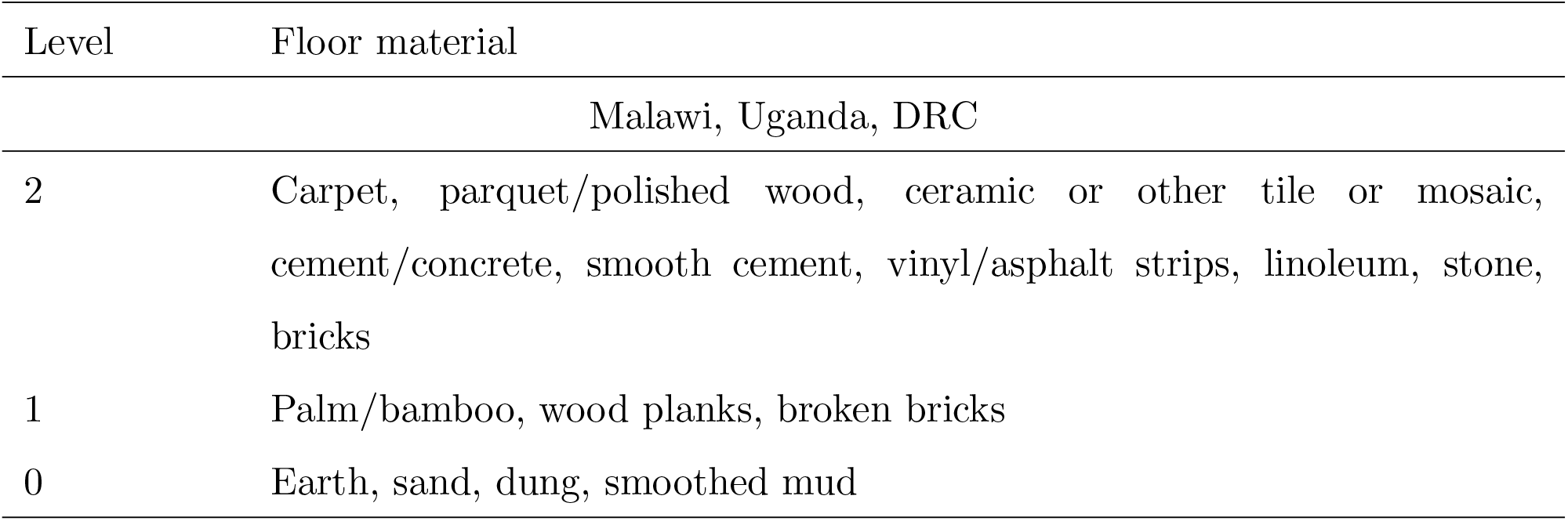
Levels for dwelling floor

| Level | Floor material |
| --- | --- |
| Malawi, Uganda, DRC |  |
| 2 | Carpet, parquet/polished wood, ceramic or other tile or mosaic, cement/concrete, smooth cement, vinyl/asphalt strips, linoleum, stone, bricks |
| 1 | Palm/bamboo, wood planks, broken bricks |
| 0 | Earth, sand, dung, smoothed mud |

**Table 1.6:** Levels for dwelling walls

| Level | Wall material |
| --- | --- |
| Malawi, Uganda, DRC |  |
| 3 | Cement, concrete, stone with lime/cement or mud, finished/burnt brick with stone or mud, covered adobe |
| 2 | Wood, wood planks/shingles, reused wood, plywood, plastic sheet, metal sheet, corrugated iron |
| 1 | Mudbrick, unburnt bricks with plaster, cement, or mud, uncovered adobe |
| 0 | Thatch, straw, grass, cane/palm/trunks, bamboo or poles with mud, timber, mud, dirt, compacted earth, cardboard, none |

**Table 1.7:**
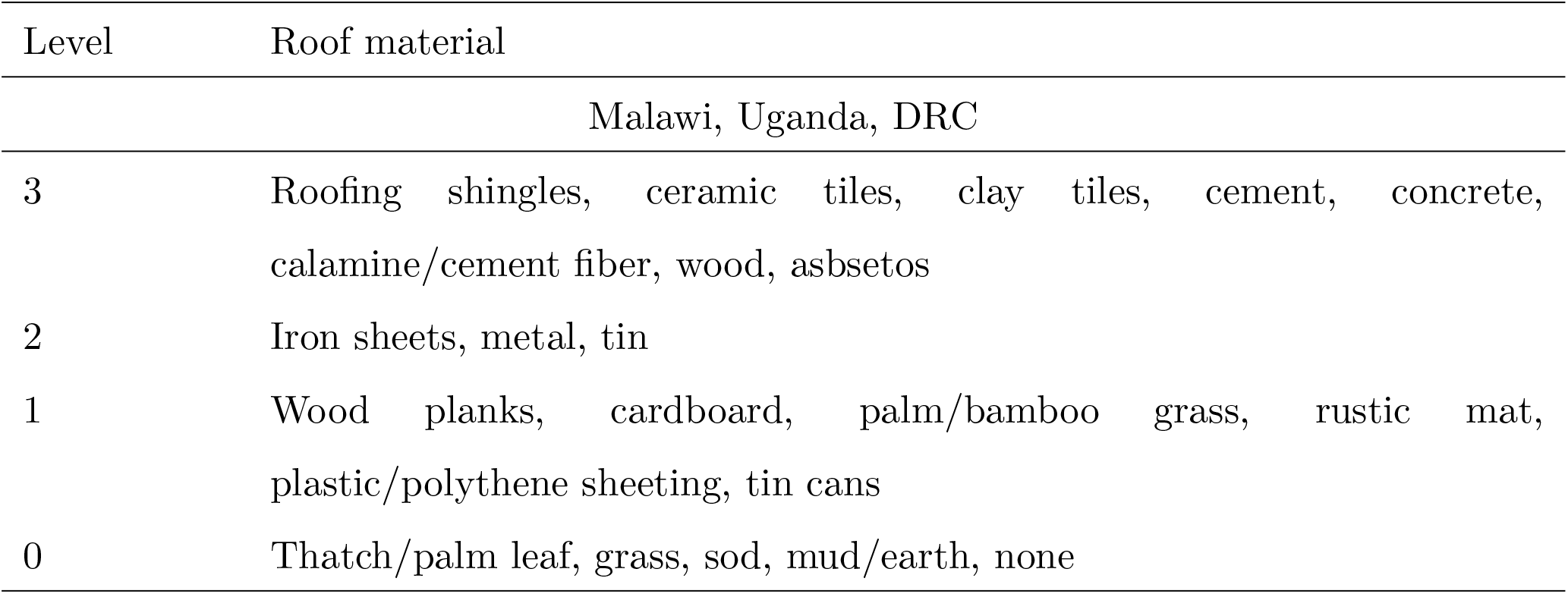
Levels for dwelling roof

| Level | Roof material |
| --- | --- |
| Malawi, Uganda, DRC |  |
| 3 | Roofing shingles, ceramic tiles, clay tiles, cement, concrete, calamine/cement fiber, wood, asbsetos |
| 2 | Iron sheets, metal, tin |
| 1 | Wood planks, cardboard, palm/bamboo grass, rustic mat, plastic/polythene sheeting, tin cans |
| 0 | Thatch/palm leaf, grass, sod, mud/earth, none |

We performed linear regression of the extracted scores on input variables as a sanity check. For all four countries, in general only variables with very large levels of missingness had negative associations with our final wealth score. In all countries, ownership of land was negatively associated with our final wealth score and had high levels of missingness (17% missing in Malawi, 25% in South Sudan, 15% in Uganda, and 31% in DRC). Similar results were found for ownership of dwelling (negative association in Malawi with 87% missing, South Sudan with 0% missing, DRC with 93% missing), area of land owned (negative association in South Sudan with 25% missing, Uganda with 92% missing, and DRC with 93% missing), and household sharing toilet with other households (negative association in Uganda with 12% missing, DRC with 15% missing). In DRC, negative associations were also found for ownership of a canoe, axe, or hoe, with 0% missing for these variables, reflecting absence of a strong association between ownership of these items and wealth. All other variables had positive associations with the final wealth score.

As the goal was to adjust for wealth score in final regression models, we did not perform factor analysis separately for urban and rural clusters in Malawi, Uganda, or DRC, however we use urban/rural status as a predictor in our models (discussed in the Mapping section, below) to reflect both the expected association between this variable and wealth score, and the stratified design of the surveys we used.

### Mapping

We detail our approach to livestock mapping in Malawi, Uganda, DRC, and South Sudan using these data in an accompanying publication, available as a preprint in S1 File. This file contains details on the stochastic partial differential equations (SPDE) to Gaussian process modeling adopted in countries with point-level data (Malawi, Uganda, and DRC), and the small area estimation approach adopted in South Sudan.

### Malawi, Uganda and DRC

After the wealth index was constructed, we collapsed over cluster by taking the mean wealth score in each cluster.

Our predictors included urban/rural status to reflect sampling strategy, and nighttime lights. Data on nighttime lights came from the National Oceanographic and Atmospheric Administration’s Nighttime Lights Time Series, which are annual cloud-free composites made from archived DMSP-OLS data, available at a resolution of 30-arc-seconds [3]. These data are available from 1992-2013, as average visible lights, stable lights, and a normalized version of average lights. We used average visible lights, and for model fitting and prediction for 2014-2020 we used data from 2013.

In addition to model selection via leave one out cross-validation as detailed in S1 File, for external validation we also performed spatial regression to check for association between our final wealth index and the proportion of the population earning under $2 per day in 2010. These latter estimates are produced by WorldPop using Bayesian model-based geostatistics and household survey data from the LSMS program, and are available at a resolution of 0.00833° [4]. We found that a one unit increase in proportion earning less than $2/day (i.e., going from 0 to 1) was associated with a 0.84 lower wealth score as predicted by our final Malawi model and a 0.87 lower wealth score as predicted by our final Uganda model, indicating good agreement between our wealth maps and the WorldPop poverty maps. WorldPop does not produce these estimates for DRC.

### South Sudan

In South Sudan we produced direct estimates of county-level wealth and design-based variance using the svydesign() and svyby() functions in the survey package and household weights provided in the IPUMS extract. In contrast with our livestock mapping methodology dtailed in S1 File we did not perform smoothing as our primary goal was not to estimate wealth, thus stabilizing variance of this estimate was not critical.

## S2 Appendix: Motivation and implementation of the parametric g-formula

### Identifiability criteria

By notational convention, let *L* be a vector of measured confounders, *Y* be outcome, *A* be exposure, and *T* be time, and let overbars denote history through time *T* = *t*. The idenfiability criteria are exchangeability, positivity, and the stable unit treatment value assumption (SUTVA).

As exchangeability, also known as no exposure-outcome confounding, is rarely satisfied marginally, the weaker assumption of exchangeability conditional on the past and on *L* is typically adopted. Using counterfactual notation, this assumption is expressed as 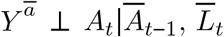.

Positivity, which refers to a positive probability of being assigned to each of the treatment levels (that is, no structural zeroes) is expressed as *Pr*(*A* = *a*|*L* = *l*) > 0 for *Pr*(*L* = *l*)≠ 0.

SUTVA has two components: consistency and no interference. Under consistency, each unit of observation (here, a cluster-year) has one potential outcome for a given treatment level *A* = *a*, and therefore a clusters observed outcome equals their counterfactual outcome under their observed exposure: if *A*_*i*_ = *a*, then 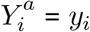 for all clusters. Under no interference, each cluster’s potential outcome is independent of all other clusters’ potential outcomes. In our study we instead assume partial interference, detailed in the main text.

### Implementation

Let *Ā* = *ā*\* be the deterministic regime under which *A* is set to *a*\* at all intervals *t* ∈ {1, …, *T*}. The g-formula is given as [1]:

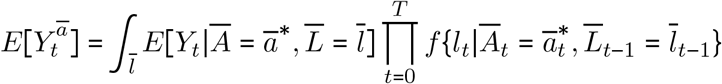

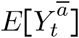 is estimated by simulating the joint distribution of 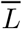 and *Y* that would have been observed had all units received exposure *Ā* = *ā* (i.e., model-based standardization). First, we defined our causal estimand *E*[*Y*^*a*^/*Y*^*a*^*], where *a*\* = empirical mean livestock density, and *a* = 1.5 × empirical mean livestock density.

Next, we examined our directed acyclic graph, contained in the main text, to identify the causal sequence of our time-varying variables, in order to write down the model for each time-varying confounder (wealth, NDVI, and LST) and the outcome (HAT cases). We handled partial interference by adjusting for mean livestock density in the interference set in the outcome (HAT) models.

Finally, we tied each of these models together by invoking the law of total probability, allowing us to obtain a marginal probability by averaging over conditional probabilities. Using the notation given above, a simplified version of the outcome model is given as:

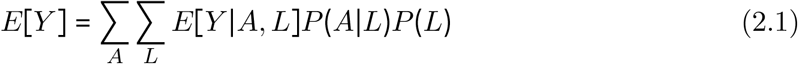

As we will implement the g-formula by setting *A* = *a*, we can remove *P (A*| *L)* from Eq 2.1. Under the identifiability criteria, we can thus identify the counterfactual parameter *E*[*Y* ^*a*^]:

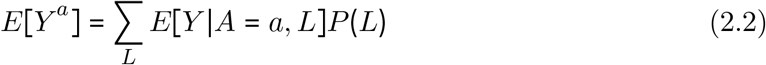

As the first term in Eq 2.2 corresponds to our outcome regression model, we need to marginalize over *P (L)* to get *E [Y* ^*a*^ ]. We do this by modeling the joint distribution of all confounders (time-varying and fixed) by taking 10,000 Monte Carlo samples from the observed data and using these samples for prediction (detailed below), ensuring all clusters which reported cases were retained. We used non-parametric bootstrapping (100 bootstrapped samples) to obtain confidence intervals.

We implemented all models using the glm() function in R. We did not use spatial models due to computational challenges and concerns regarding spatial confounding. We fit Gaussian models for wealth, NDVI, and LST, and Poisson models for HAT cases. Our procedure was as follows, iterating over each *t* with *t*_0_ defined as 2001 (2000 was not modeled due to the need to implement lags):

1. For *t*_0_, fit all models except the outcome model (HAT cases): wealth, NDVI, and LST. Outcome model not fit due to the need to lag livestock by two time points (years)
2. Set livestock density to *A* = *a*, and for each model in turn use the predict() function to predict the variable in question under *A* = *a* on the Monte Carlo sample
3. Repeat for all times *t*_1_, …, *t*_*T*_, adding in the outcome model for *t* > 0.
4. Iterate steps 1-3 over each bootstrapped sample

The result of this procedure is a vector of counterfactual outcomes for each simulation *s*, with 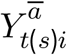 defined as the 1-year cumulative incidence of HAT in cluster *i*, year *t*, simulation *s*, and with exposure set to *Ā* = *ā*.

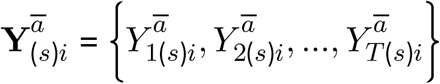

As noted above, we take the ratio of the counterfactual outcomes under *Ā* = *ā* versus under *Ā =ā*\* as the target of inference. Across clusters, time and simulations, we implement this as follows, separately for each country (and for gHAT and rHAT in Uganda):

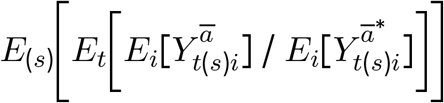

with uncertainty bounds estimated by taking the 2.5% and 97.5% quantiles across simulations.

## S3 Appendix: Implementation of the measurement error model in South Sudan

The measurement error (MEC) model is defined as:

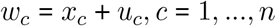

where *c* indexes county, *u*_*c*_ are the measurement error terms distributed as 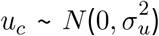, and *x*_*c*_ and *u*_*c*_ are independent. Since the *u* terms have mean 0, *E (W*| *X* = *x)* = *x*, that is, *W* is unbiased for a given unobserved *x*. Failing to account for measurement error will result in biased effect estimates and inappropriate standard errors [1].

Following the heteroscedastic errors-in-variables approach detailed in Wang et al. (2018), we defined our hierarchical model as follows:

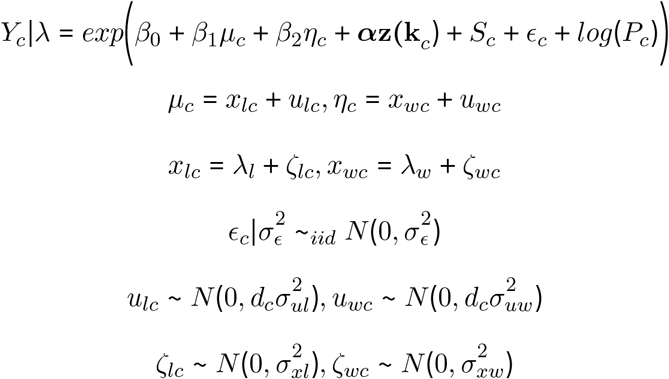

where

- *Y*_*c*_ is the number of cases in county *c*
- *µ*_*c*_ is estimated livestock (cattle or pig) density in county *c*
- *η*_*c*_ is estimated wealth score in county *c*
- ***α*** is a vector of coefficients
- **z(k**_*c*_) is a vector of confounders measured without error
- *P*_*c*_ is the offset, given as population in county *c*
- *ϵ*_*c*_ are county-level iid (unstructured) random effects with variance 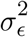
- *S*_*c*_ are county-level structured random effects which follow the ICAR model with marginal variance 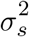
- *ne*(*c*) denotes neighbors (shared boundary) of county *c*
- *m*_*c*_ is the number of neighbors of county *c*
- *x*_*lc*_ is the true livestock density in county *c*
- *u*_*lc*_ is the measurement error for livestock density in county *c*
- *x*_*wc*_ is the true wealth score in county *c*
- *u*_*wc*_ is the measurement error for wealth score in county *c*
- *λ*_*l*_ is the mean of the true livestock density
- *λ*_*w*_ is the mean of the true wealth score
- *ζ*_*lc*_ is the residual for livestock density in county *c*
- *ζ*_*wc*_ is the residual for wealth score in county *c*
- *d*_*c*_ is a weight which allows for heteroscedasticity in the error structure

For both livestock density and wealth score, we assume 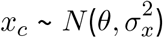, where *θ* is the mean of *x*_*c*_ and 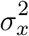 is the variance.

There are therefore four parameters needed for each of the two resulting measurement error models: *β*_1_ (*β*_2_ for wealth score), 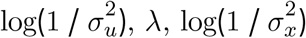, as well as the scale factor *d*_*c*_. For livestock density, we specified the priors and starting values for these parameters as follows for livestock, substituting *µ* for *η* and *β*_1_ for *β*_2_ for wealth:

- 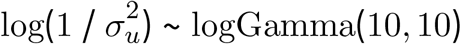, starting value log(1 / *var*(*σ*_*µ*_))
- *β*_1_ ∼ Normal 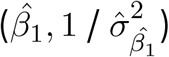.
- *λ* fixed at mean *E*[*µ*], with a Gaussian prior
- 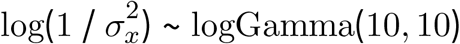, starting value log(1 / *var*(*µ*))
- *d*_*c*_ ∼ Unif(0.5, 1.5)

where *var*(*σ*_*µ*_) is the empirical variance of the posterior standard deviation for livestock density, 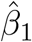 is the posterior mean of the coefficient for livestock density from the naive model, 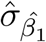 is the posterior standard deviation of the same coefficient from the naive model, and *var*(*µ*) is the empirical variance of the posterior median for livestock density. The logGamma prior specification is equivalent to that used in Wang et al. [1]. The MEC model for wealth was specified equivalently, and in our final models both livestock density and wealth were included in this form.

## S4 Appendix: Descriptive statistics plots

### Malawi

**Figure 4.1:**
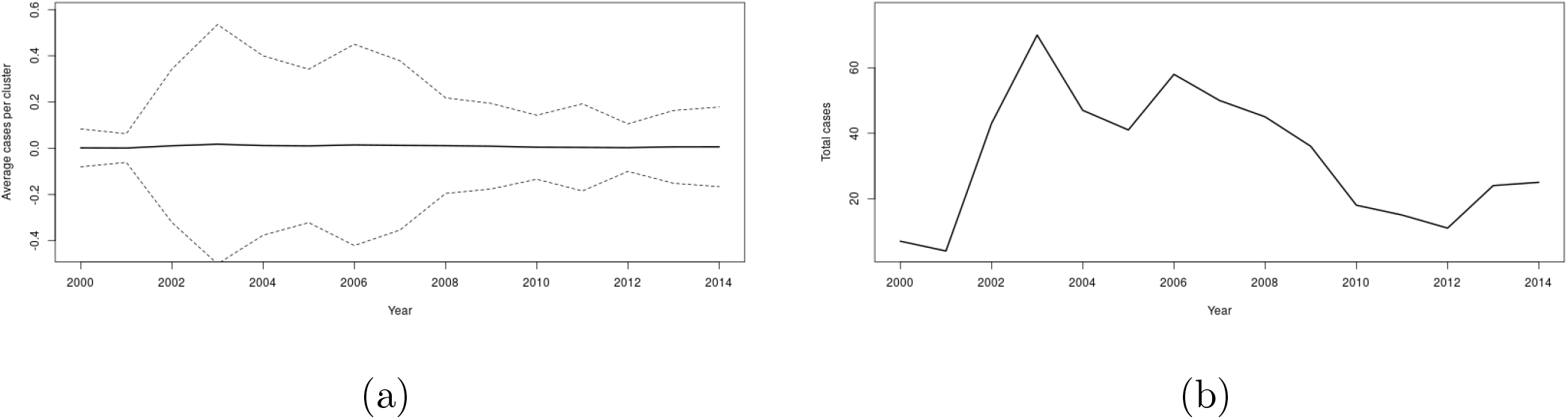
Mean (a) and sum (b) of cases over time in study clusters, Malawi

**Figure 4.2:**
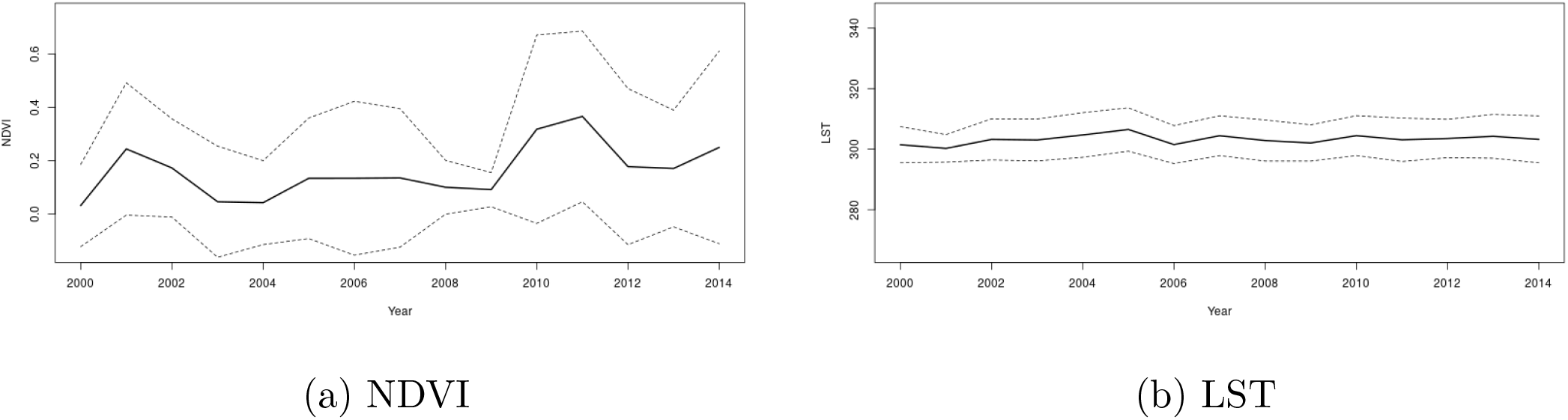
NDVI (a) and LST (b) over time in study clusters, Malawi

**Figure 4.3:**
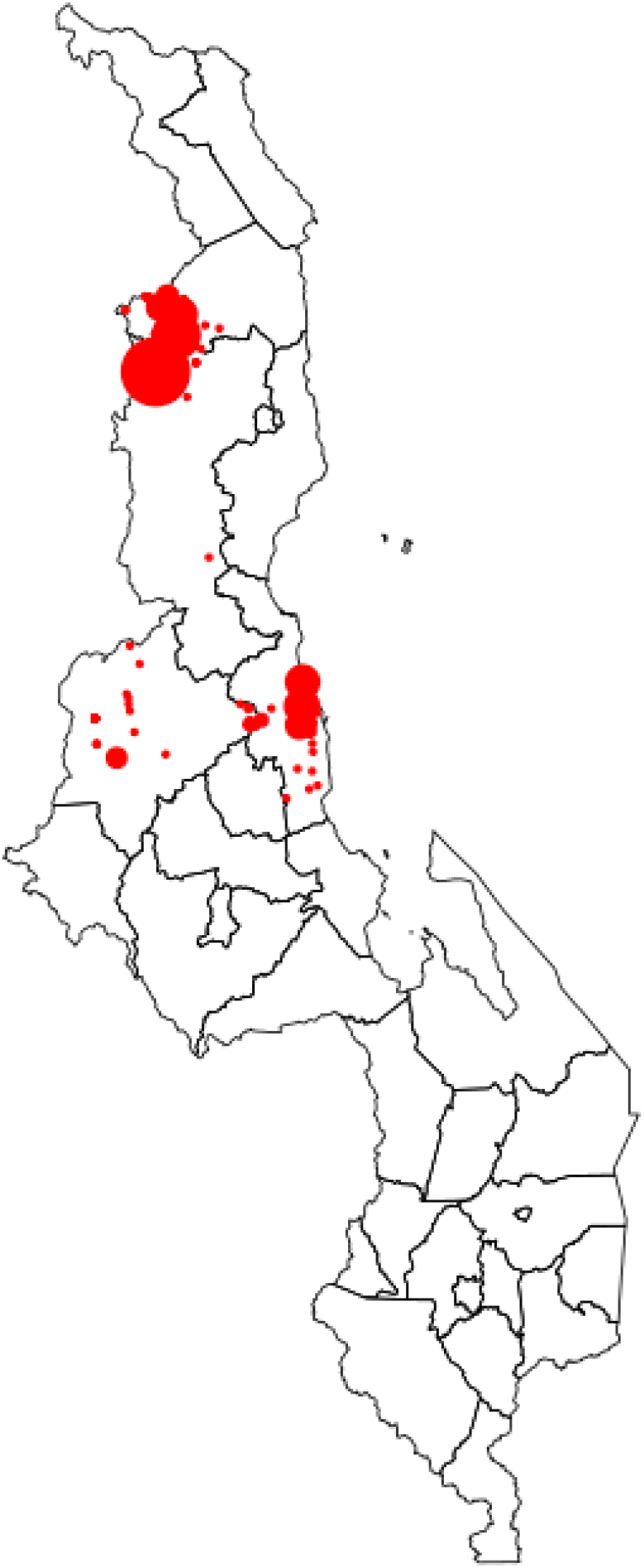
HAT cases 2000-2014, Malawi

**Figure 4.4:**
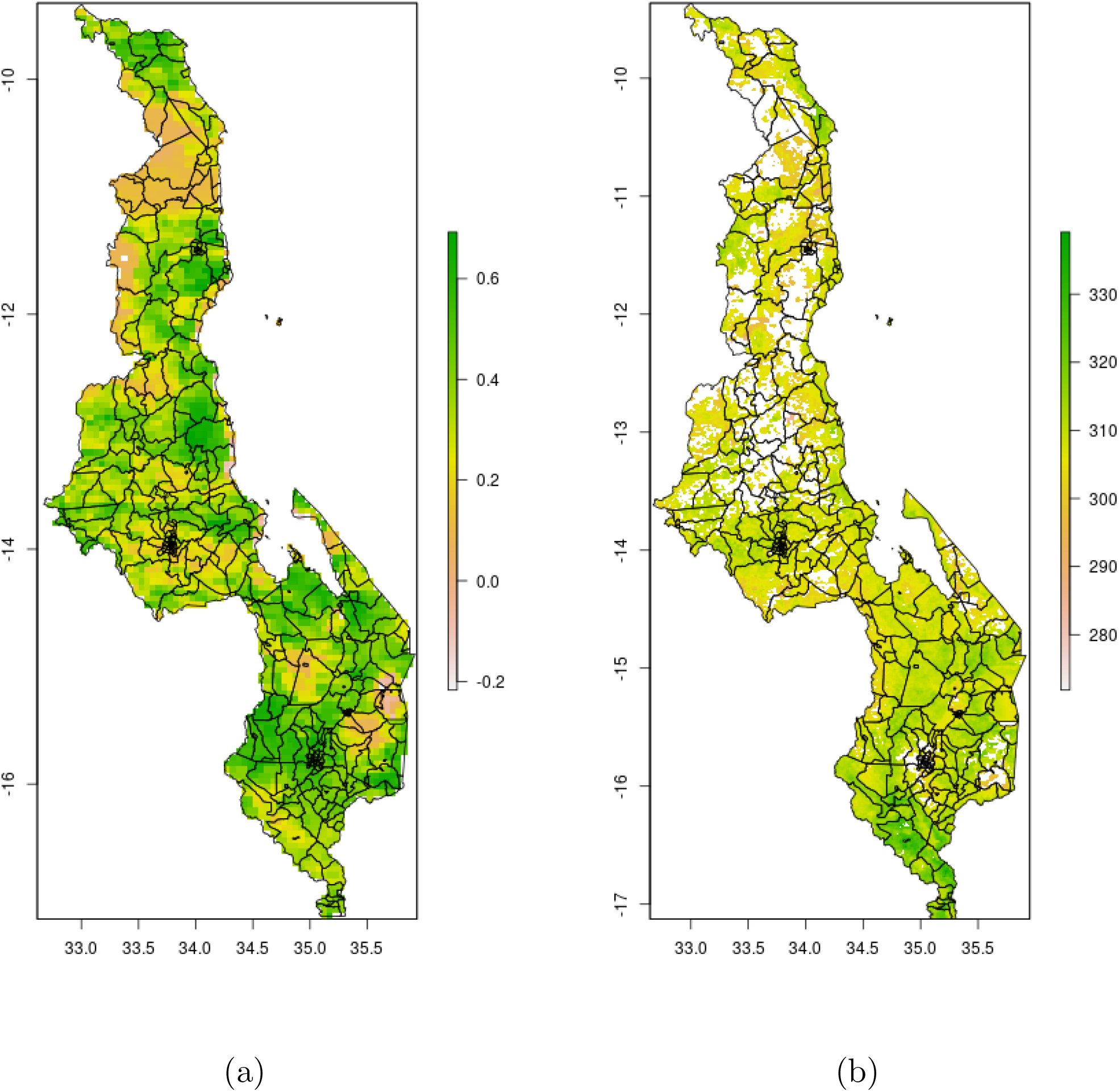
2010 NDVI (a) and LST (b), Malawi

**Figure 4.5:**
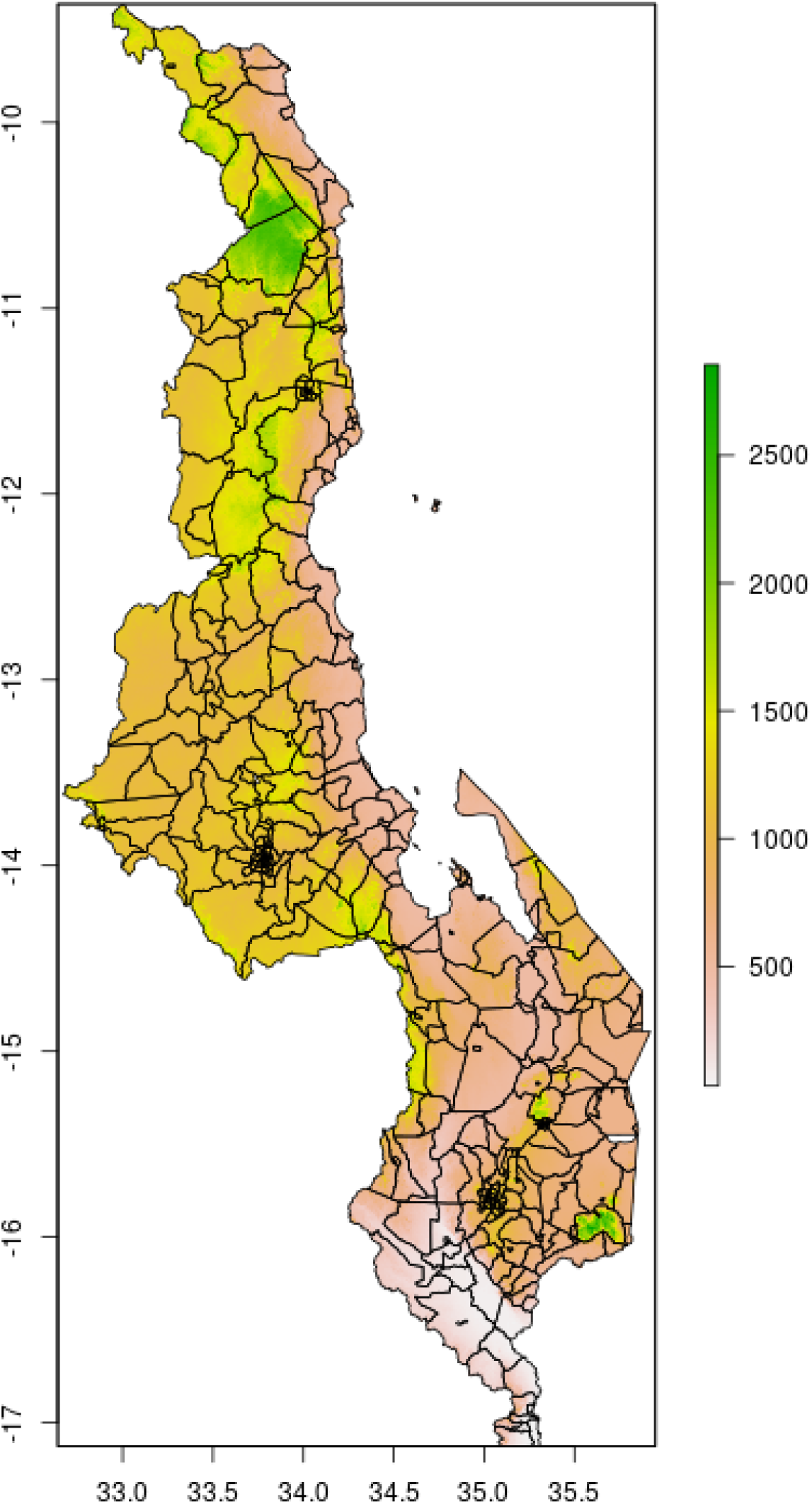
Elevation, Malawi

**Figure 4.6:**
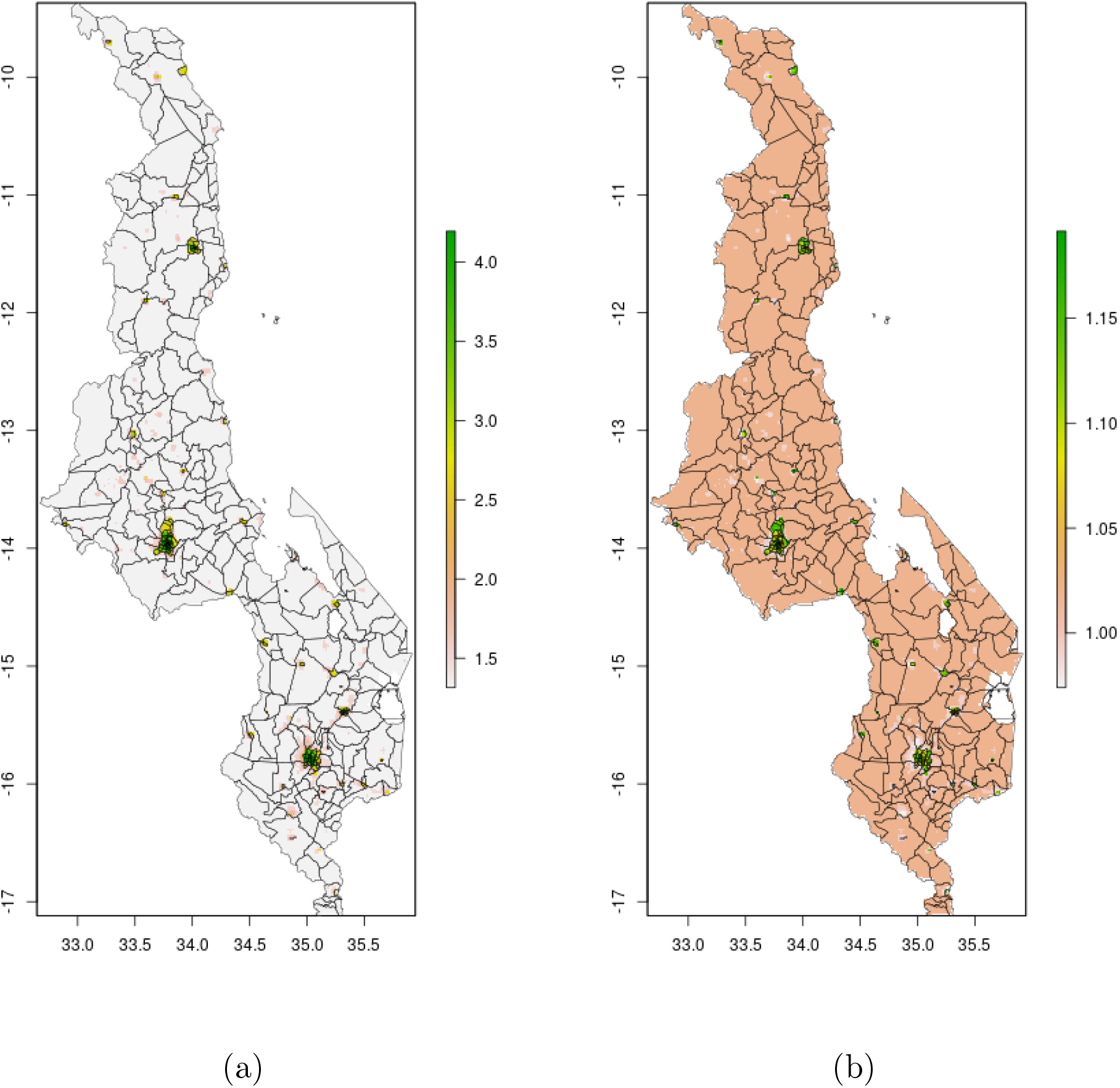
2010 wealth scores, mean (a) and posterior 95% credible interval (b), Malawi

### Uganda

**Figure 4.7:**
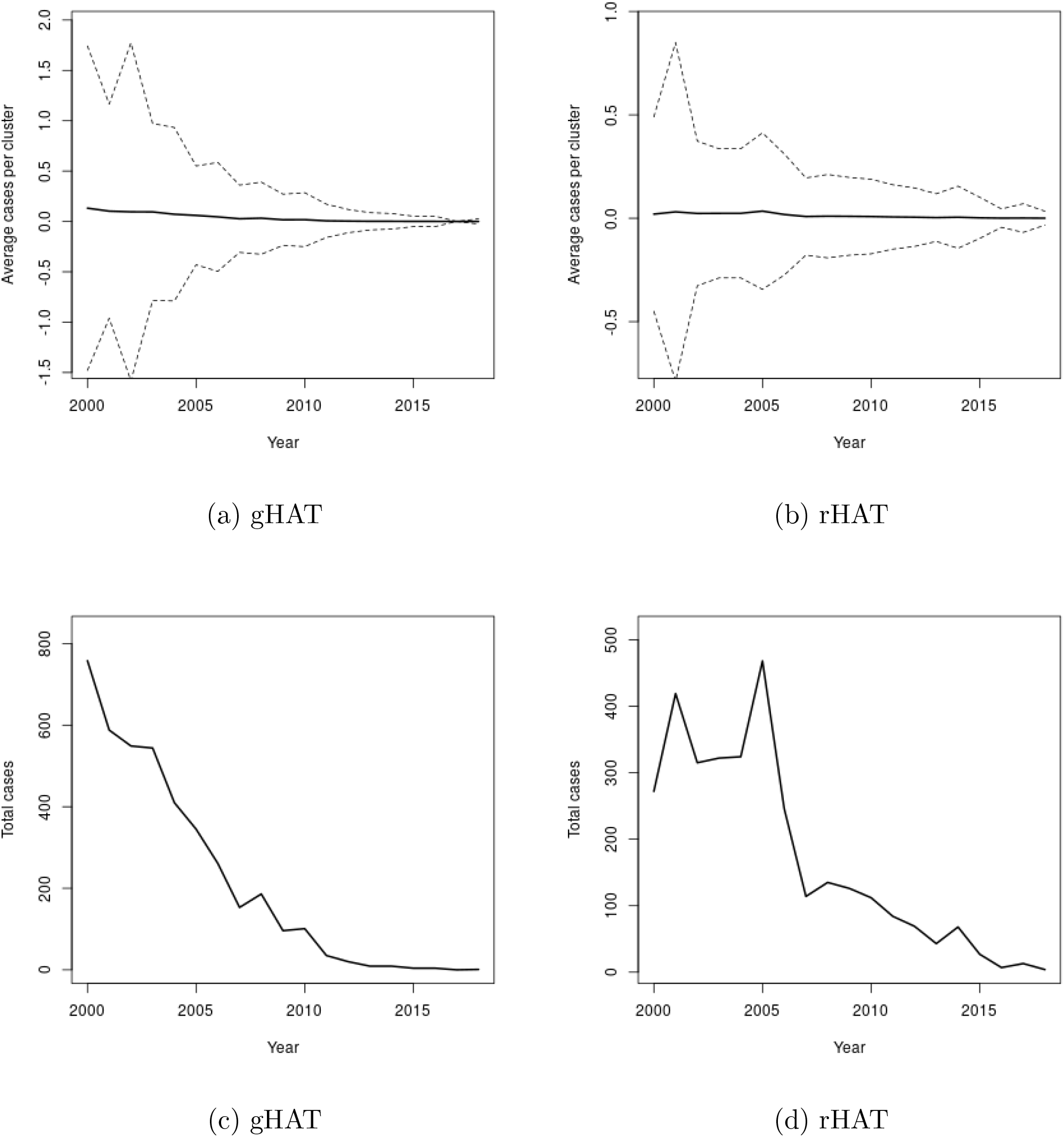
Mean (a-b) and (c-d) of cases over time in study clusters, Uganda

**Figure 4.8:**
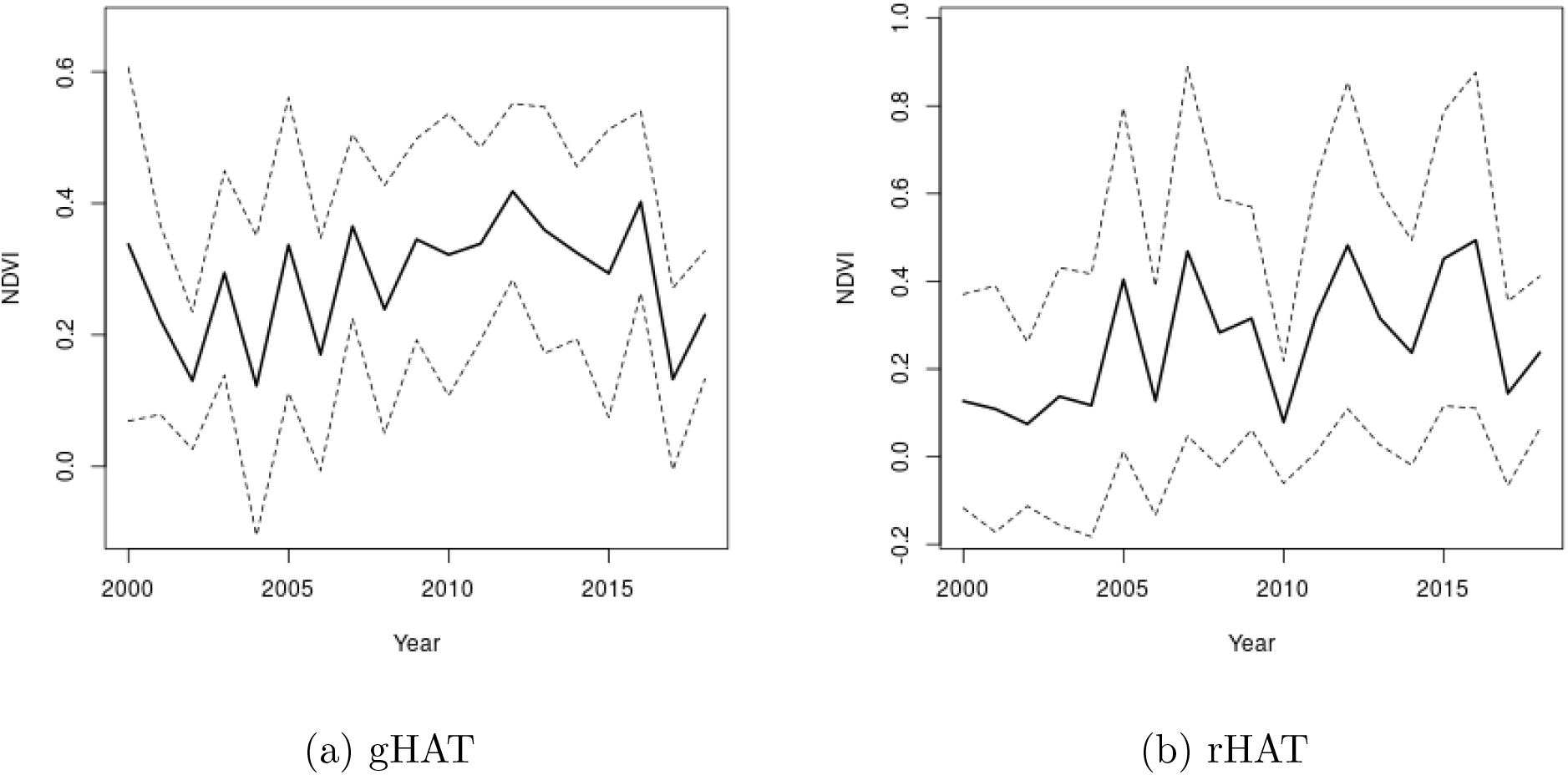
NDVI over time in study clusters, Uganda

**Figure 4.9:**
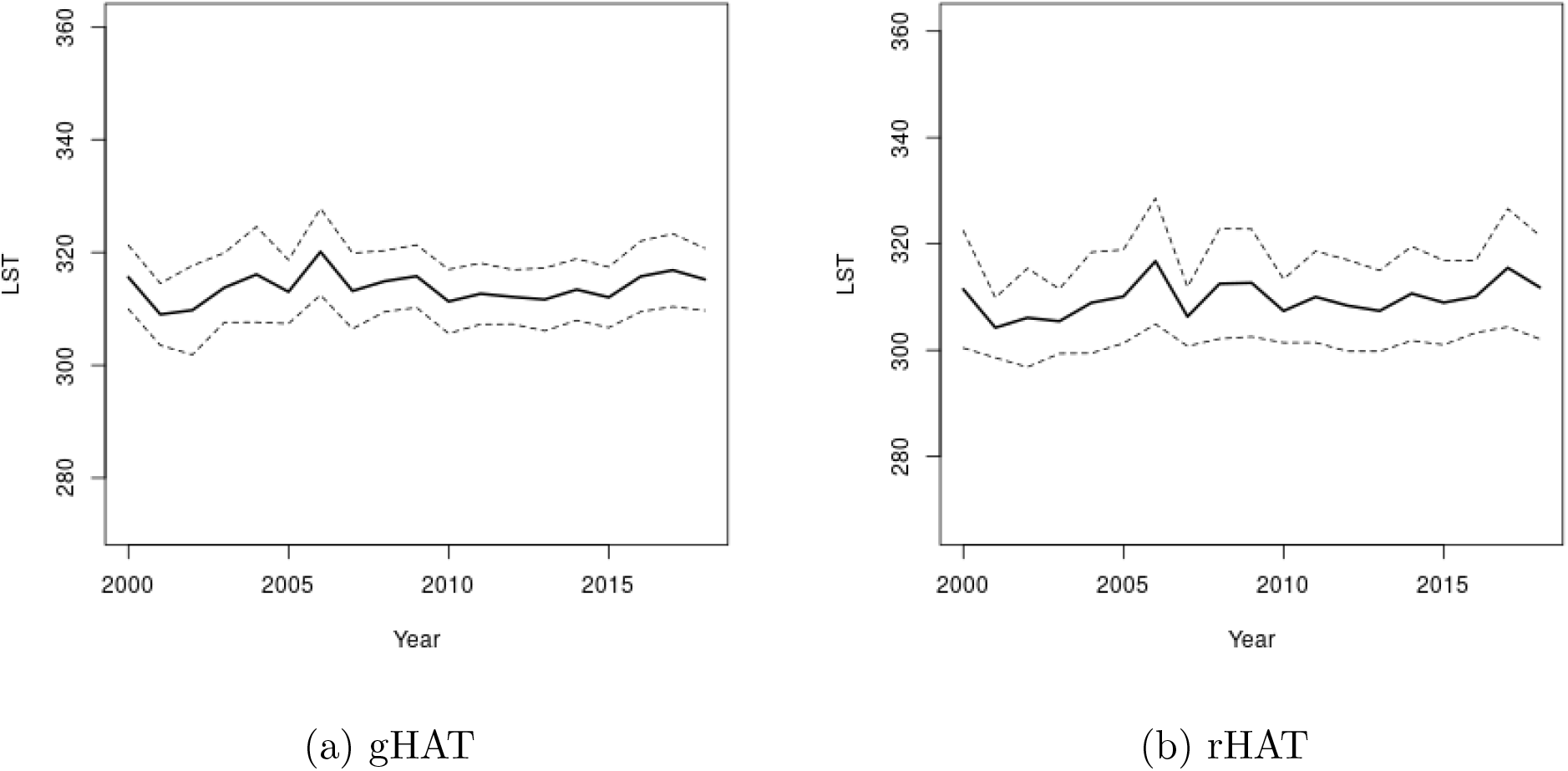
LST over time in study clusters, Uganda

**Figure 4.10:**
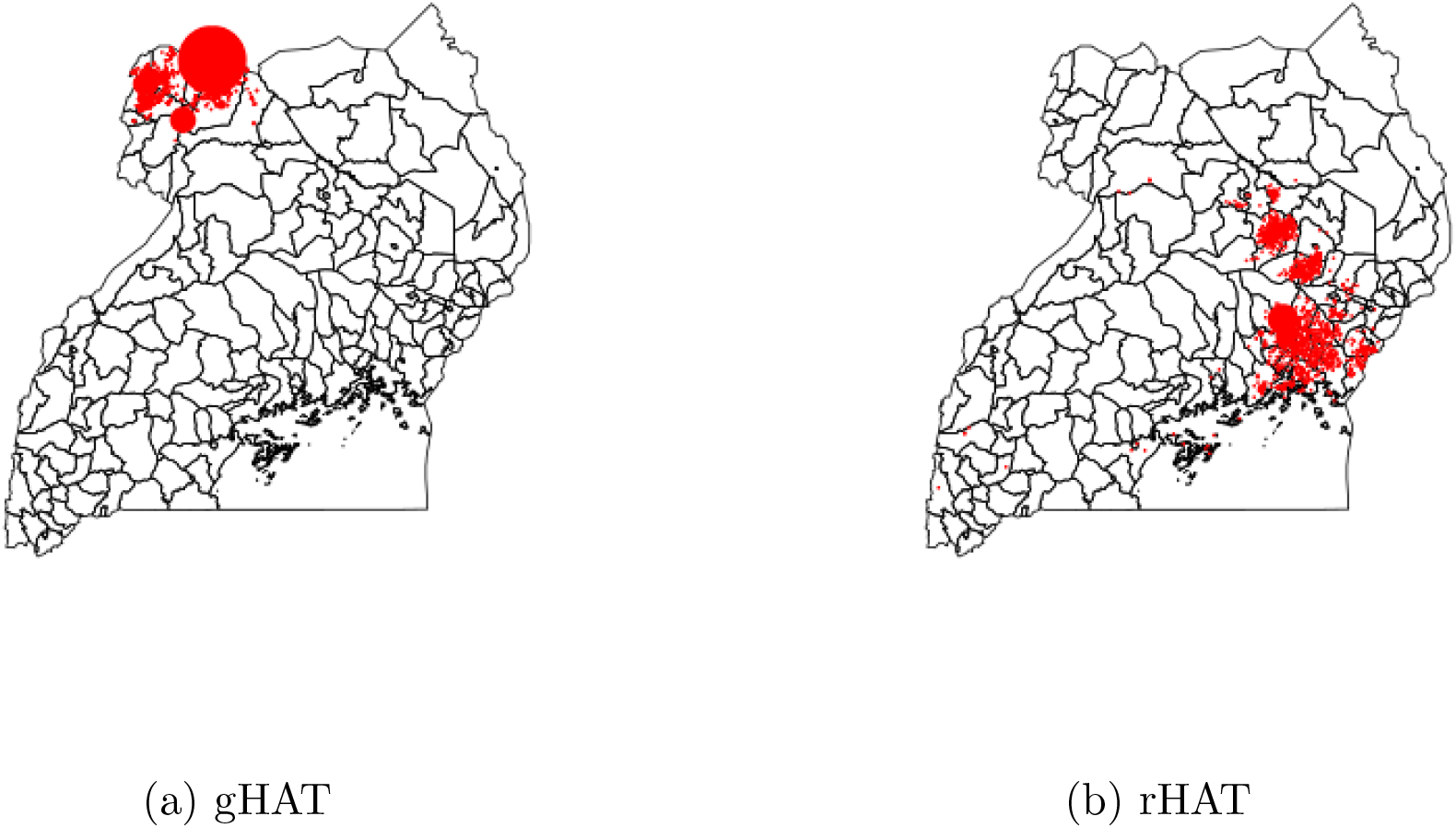
HAT cases 2000-2018, Uganda

**Figure 4.11:**
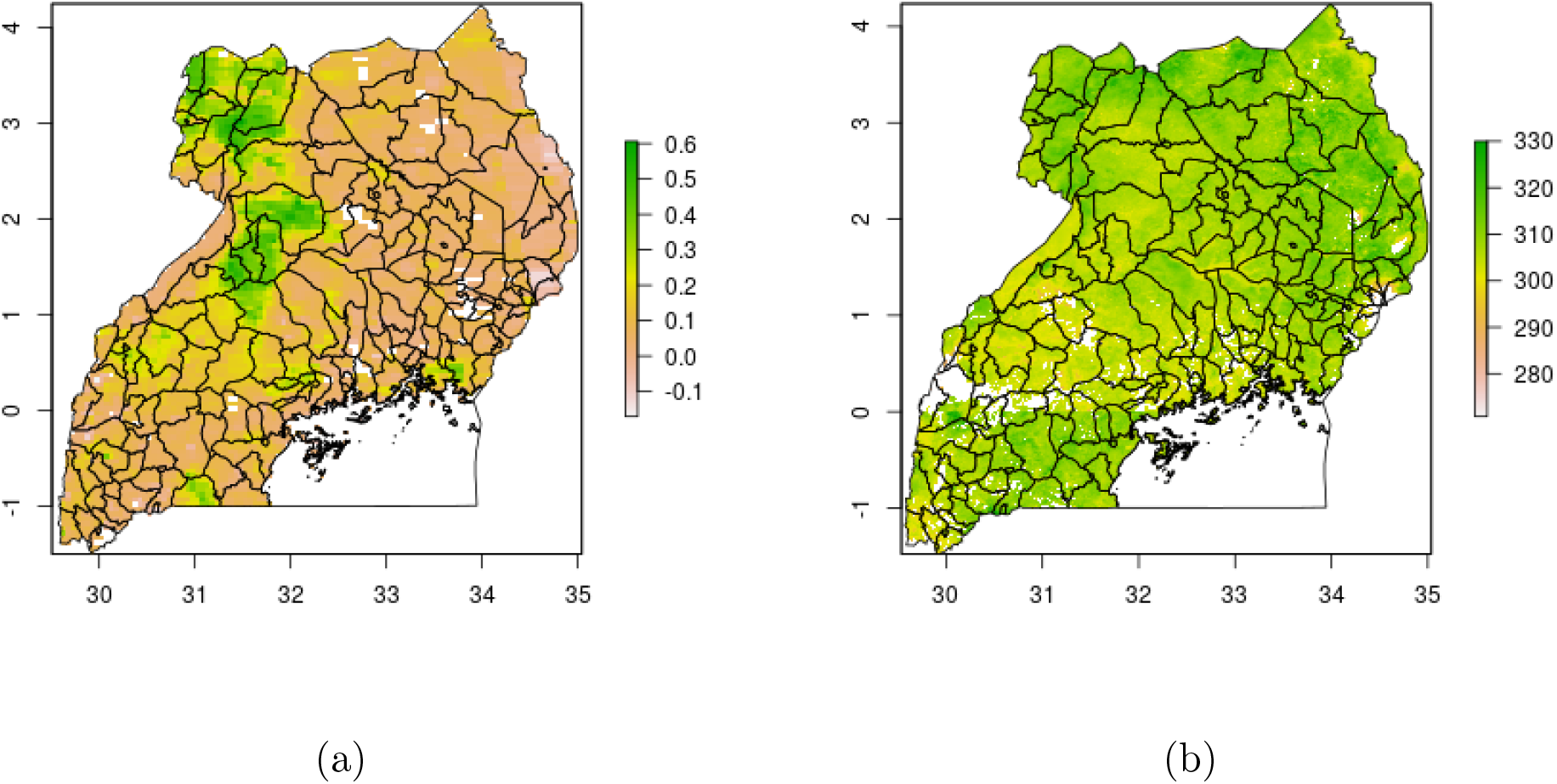
2010 NDVI (a) and LST (b), Uganda

**Figure 4.12:**
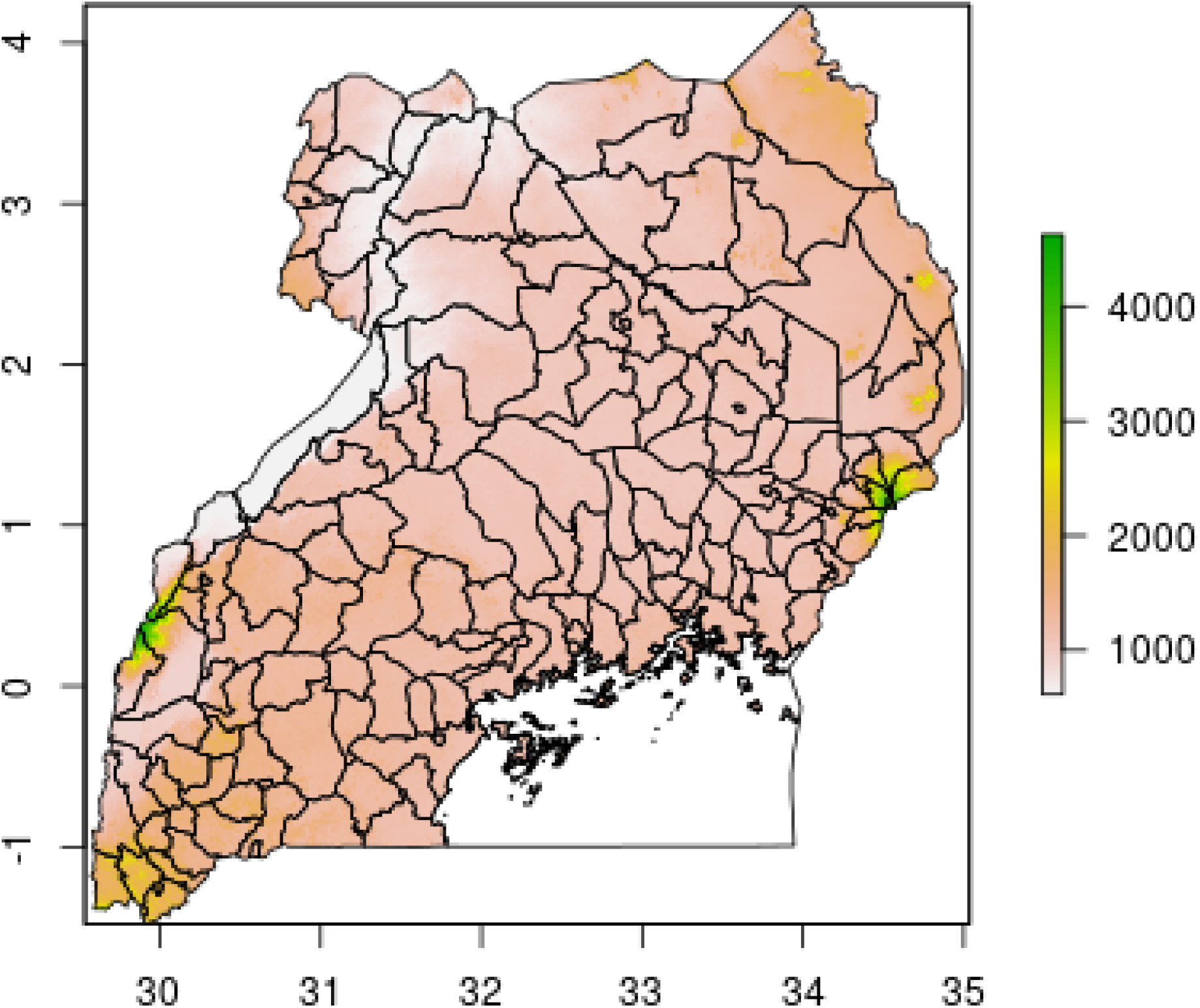
Elevation, Uganda

**Figure 4.13:**
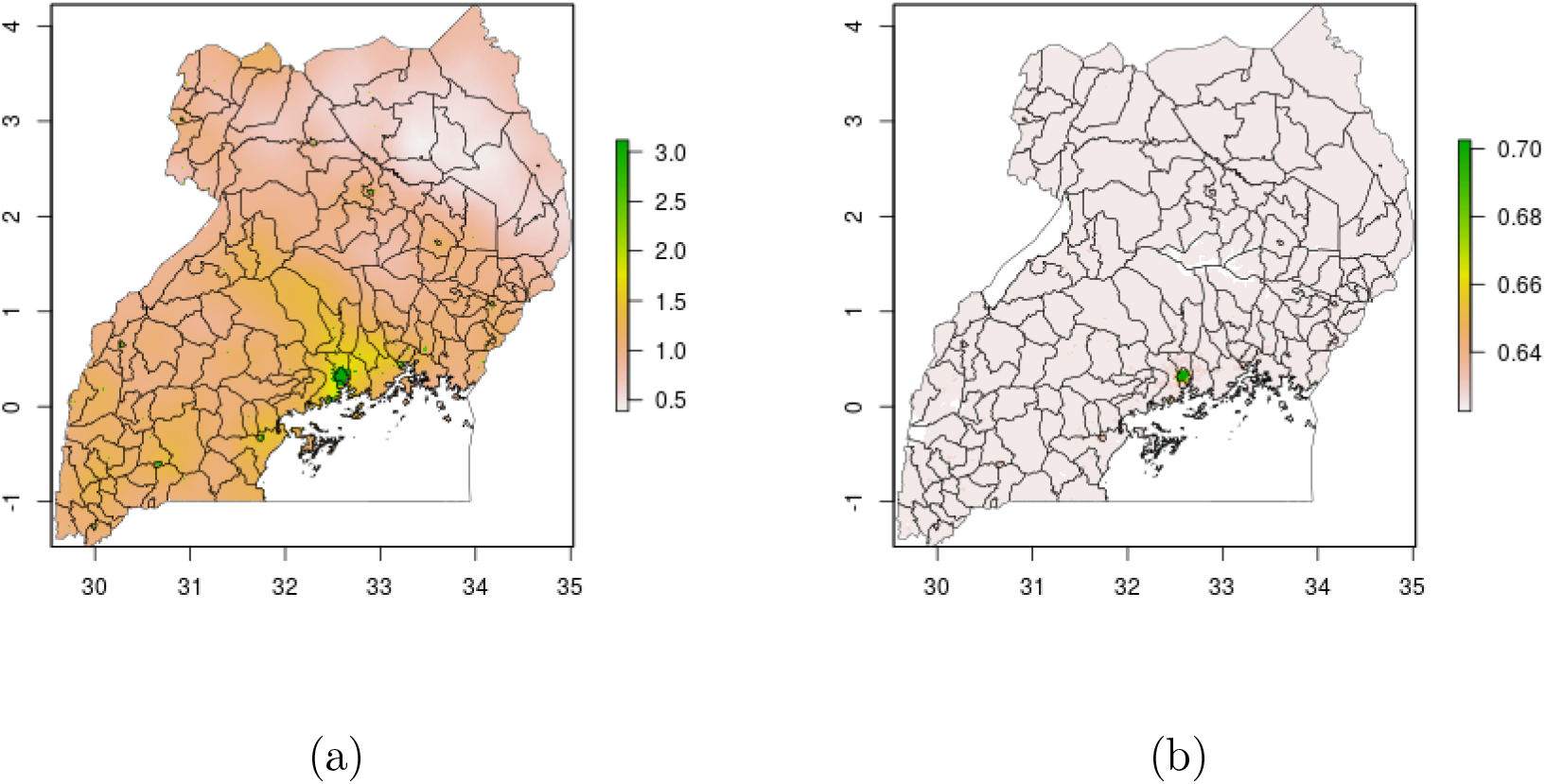
2010 wealth scores, mean (a) and posterior 95% credible interval (b), Uganda

### DRC

**Figure 4.14:**
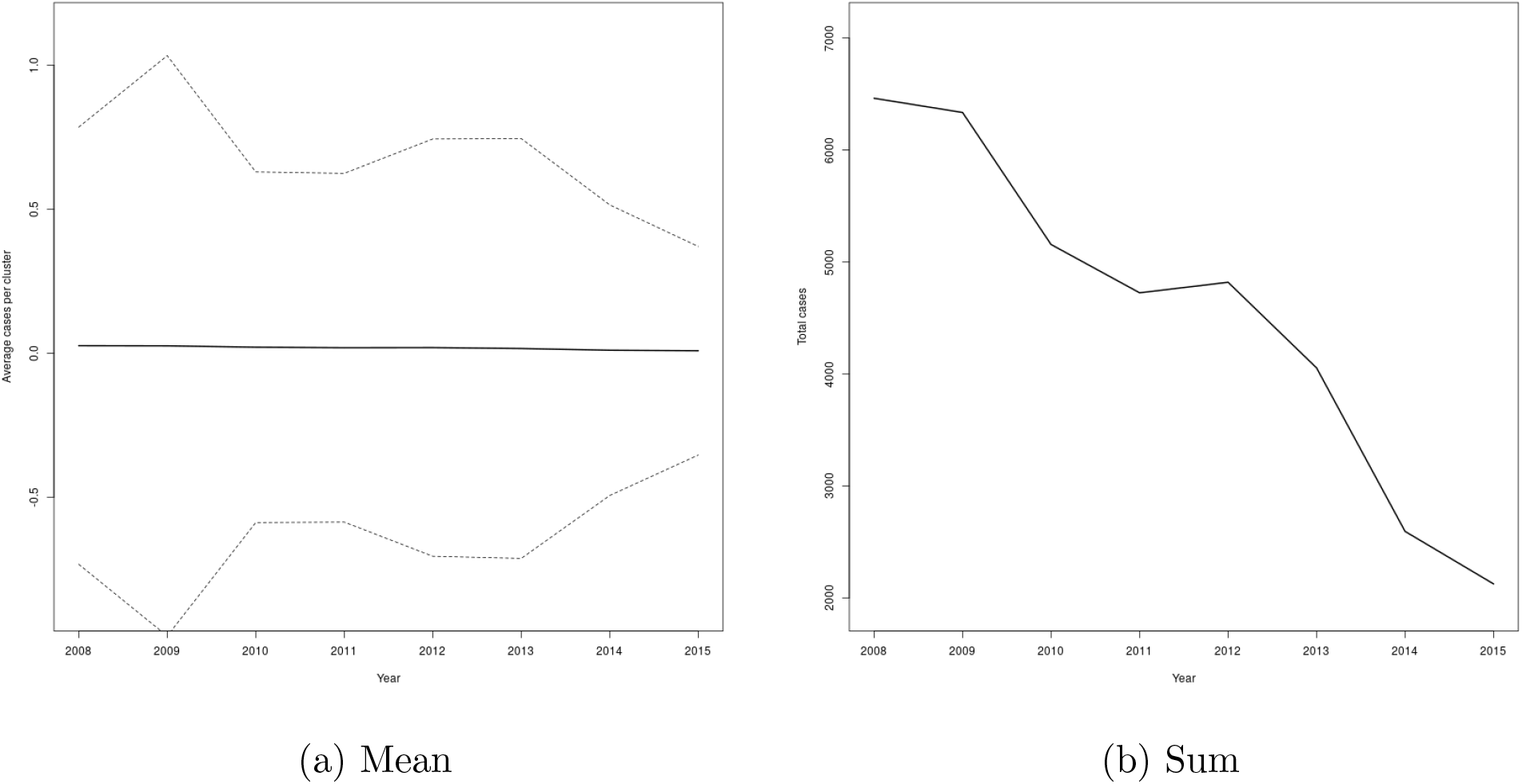
Mean (a) and sum (b) of cases over time in study clusters, DRC

**Figure 4.15:**
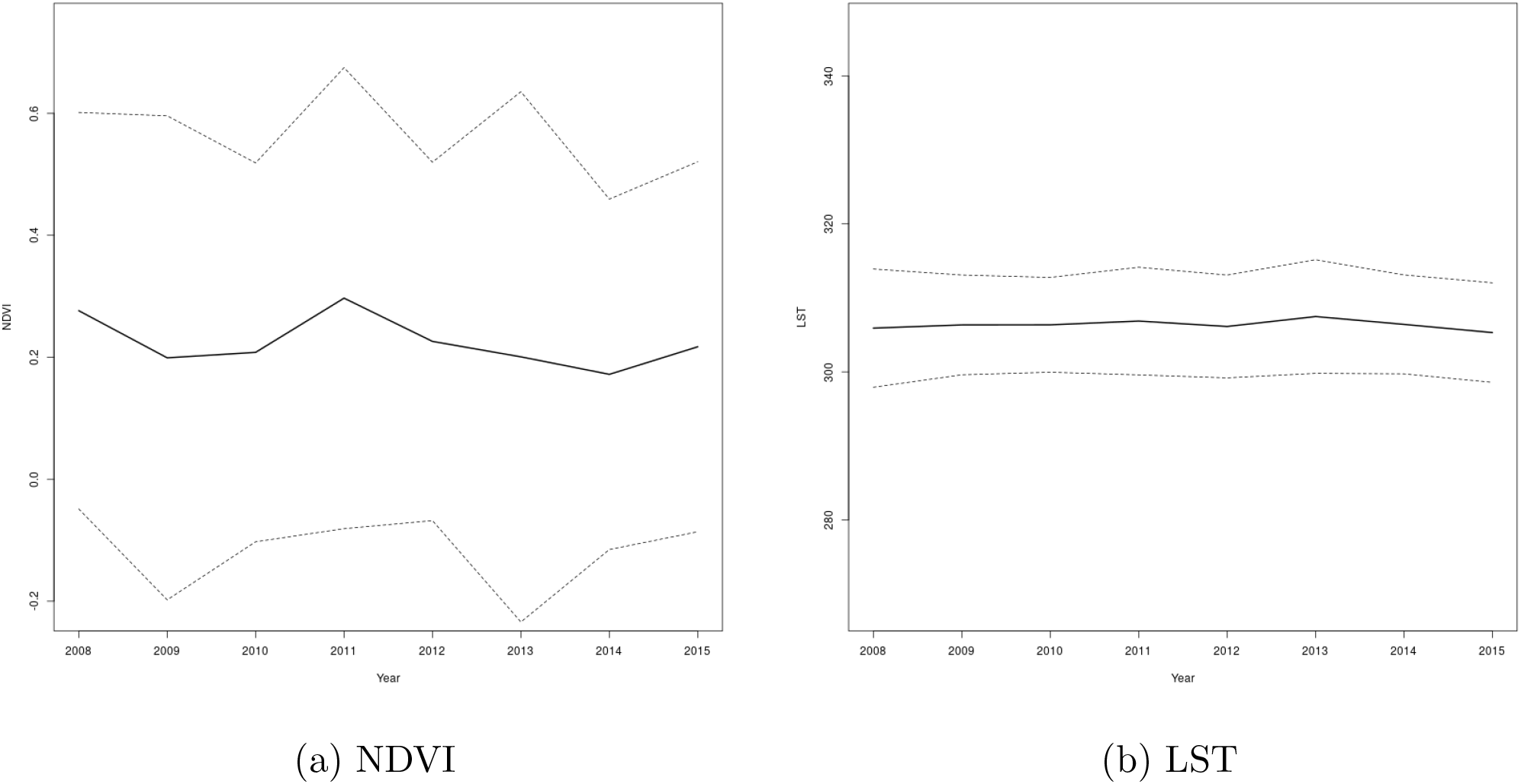
NDVI (a) and LST (b) over time in study clusters, DRC

**Figure 4.16:**
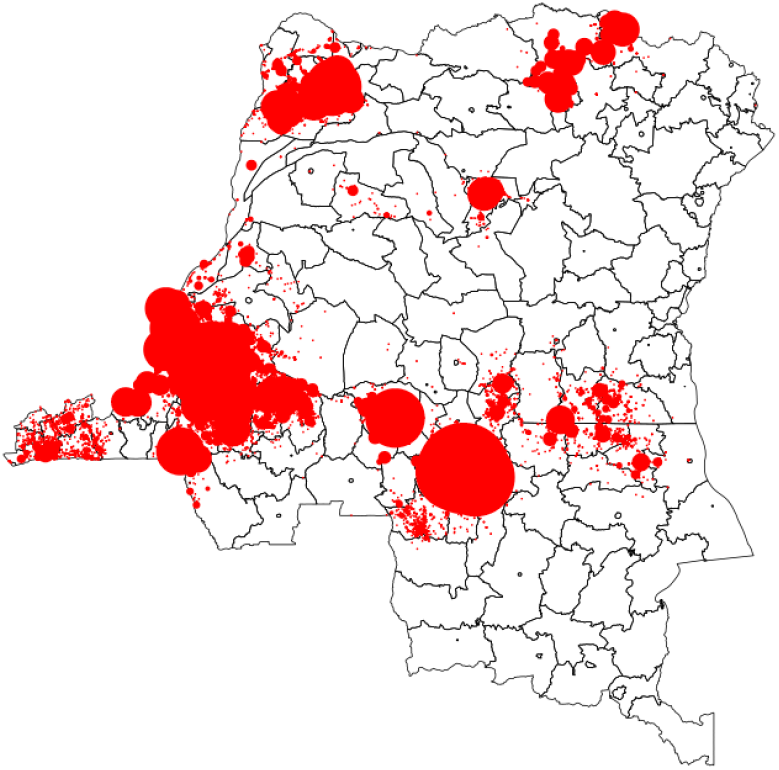
HAT cases 2008-2015, DRC

**Figure 4.17:**
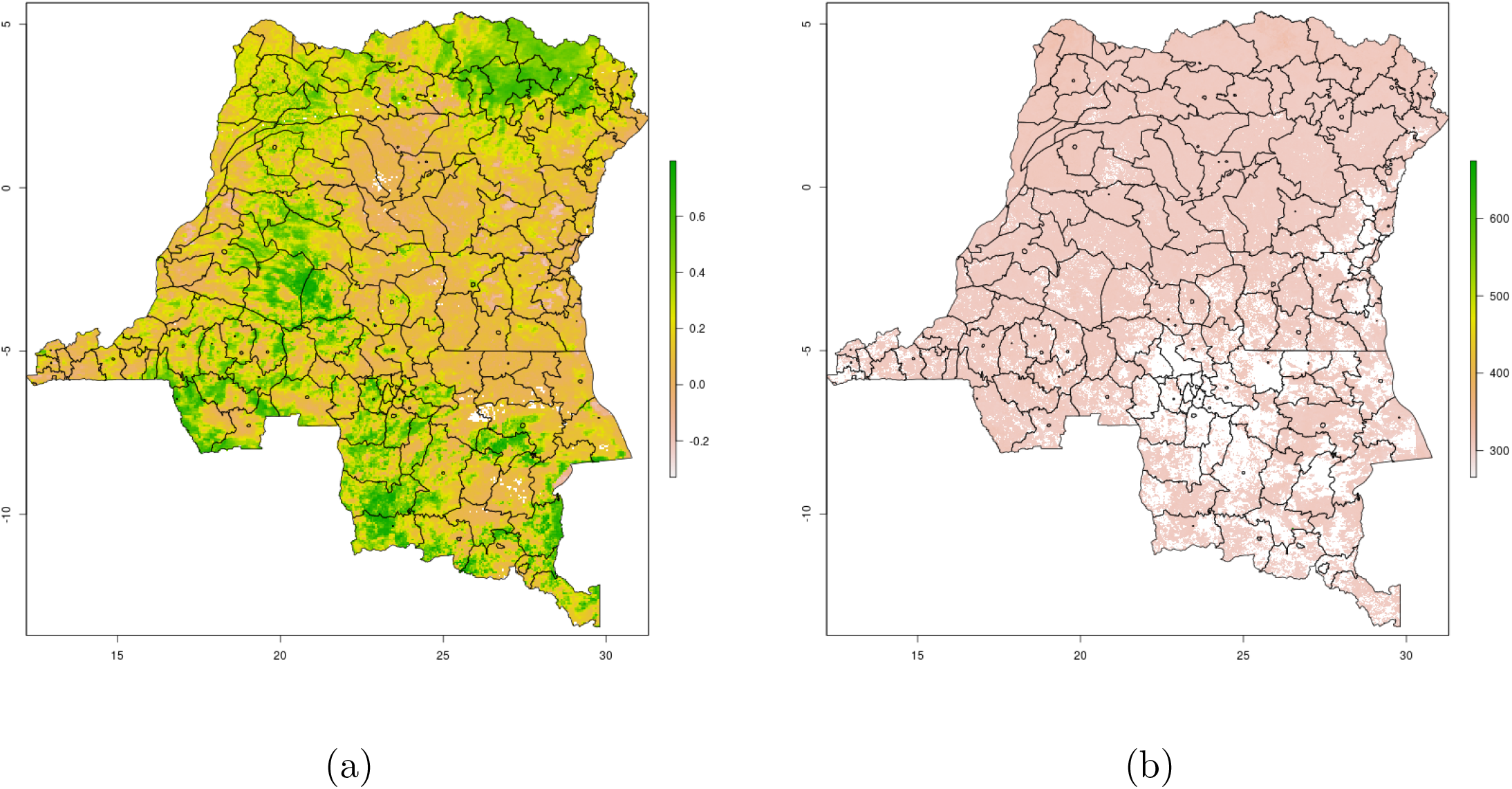
2010 NDVI (a) and LST (b), DRC

**Figure 4.18:**
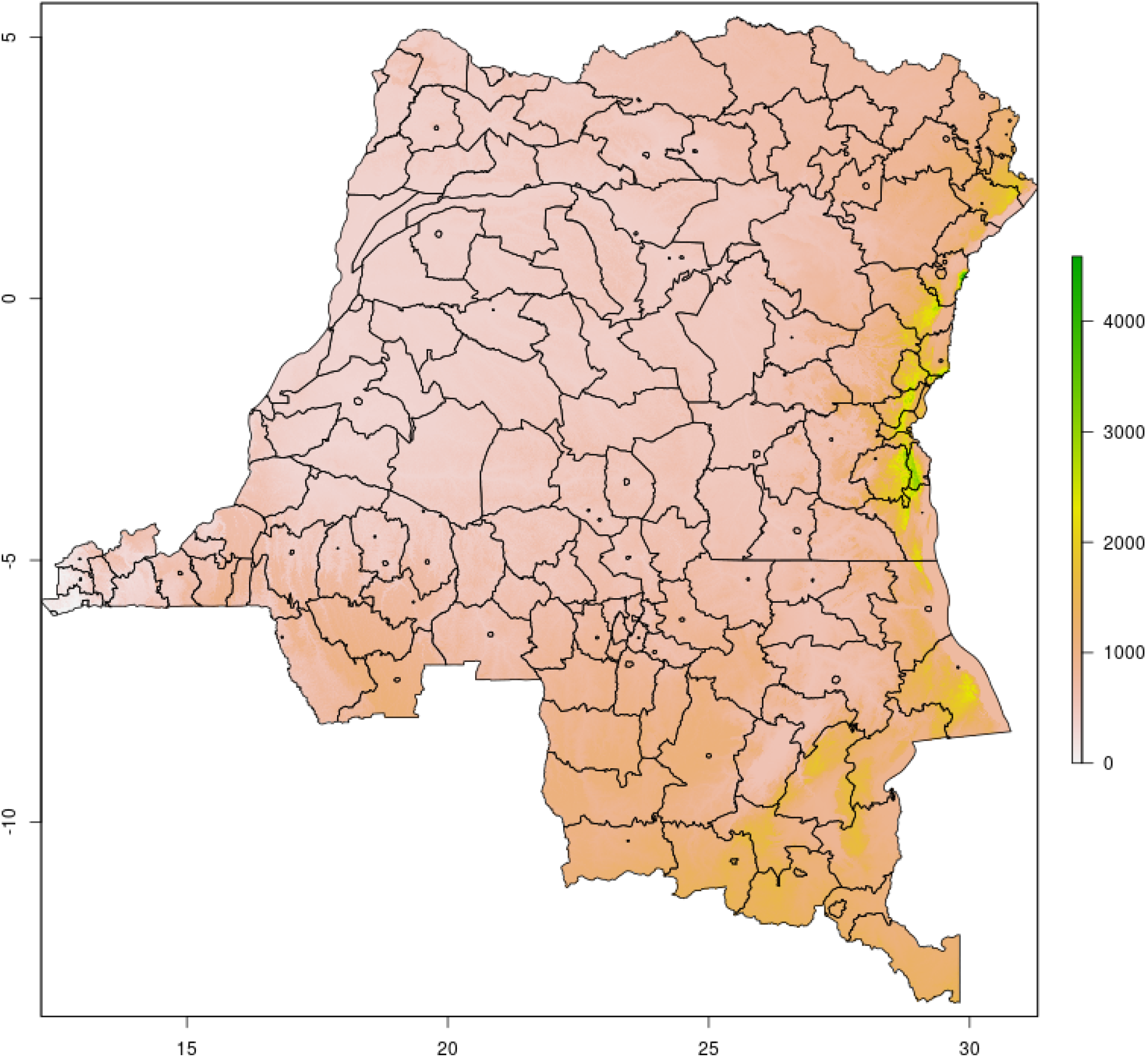
Elevation, DRC

**Figure 4.19:**
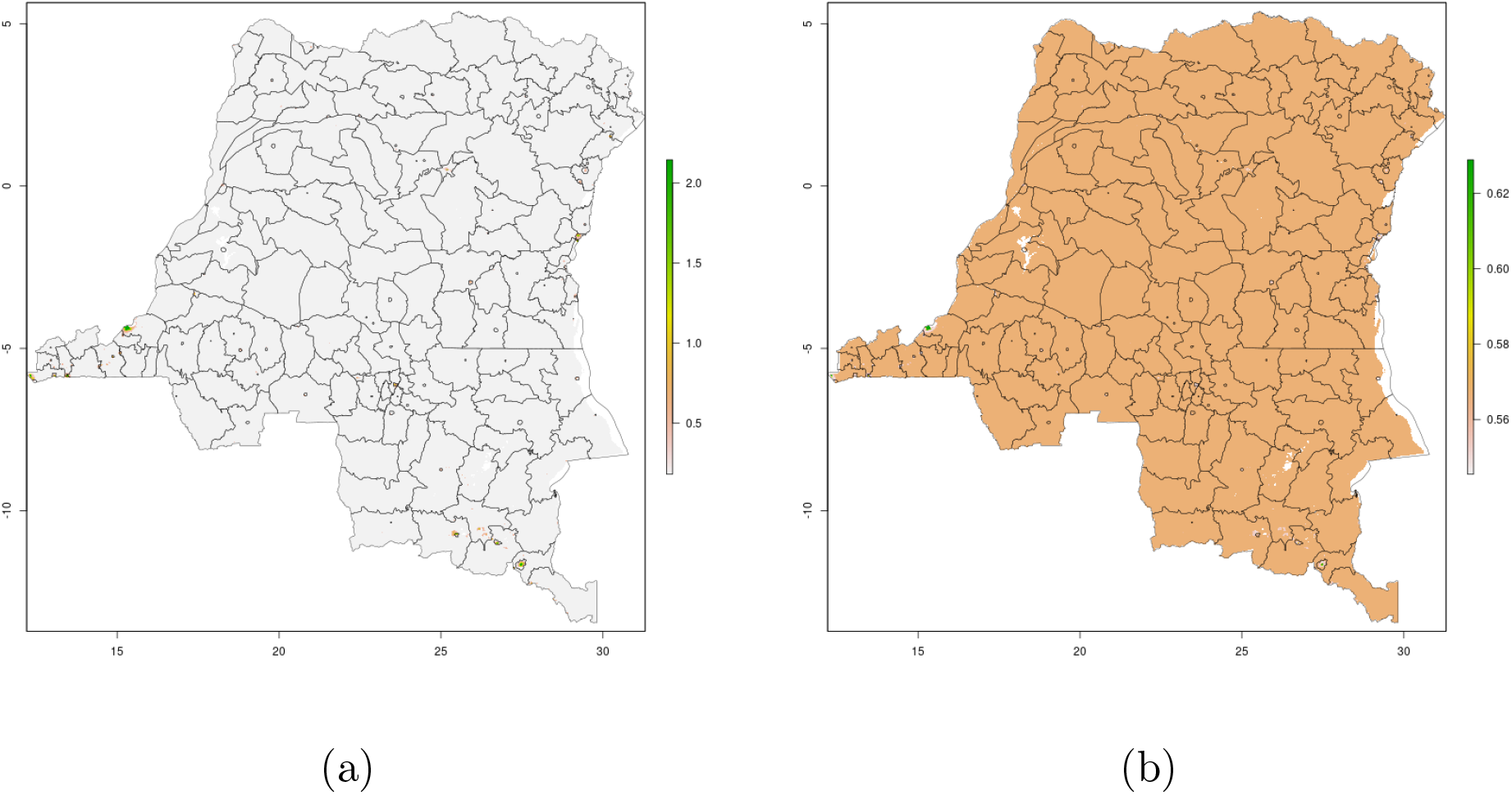
2010 wealth scores, mean (a) and posterior 95% credible interval (b), DRC

### South Sudan

**Figure 4.20:**
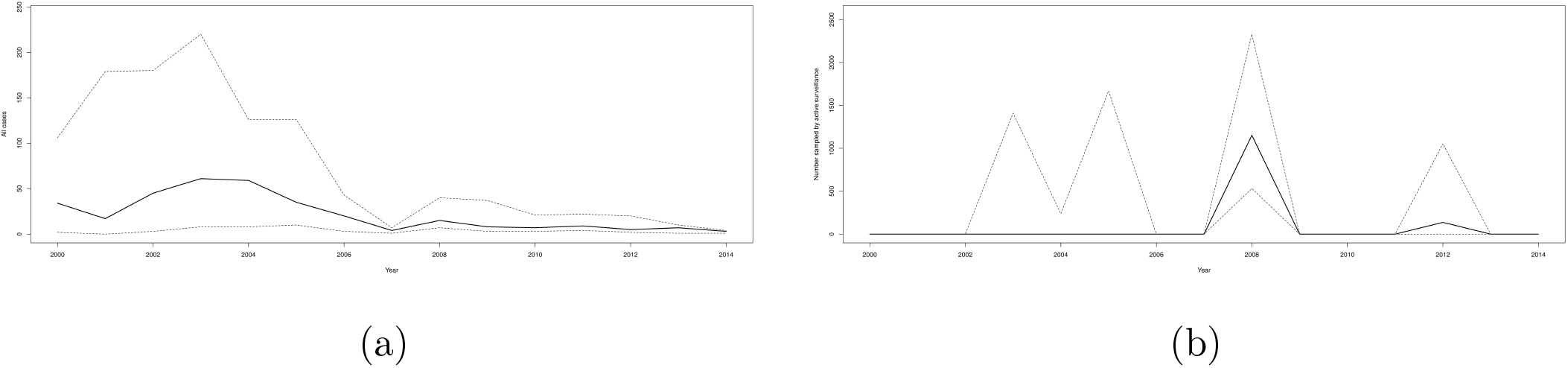
Mean number of cases (a) and people sampled by active surveillance (b) over time in study counties, South Sudan

**Figure 4.21:**
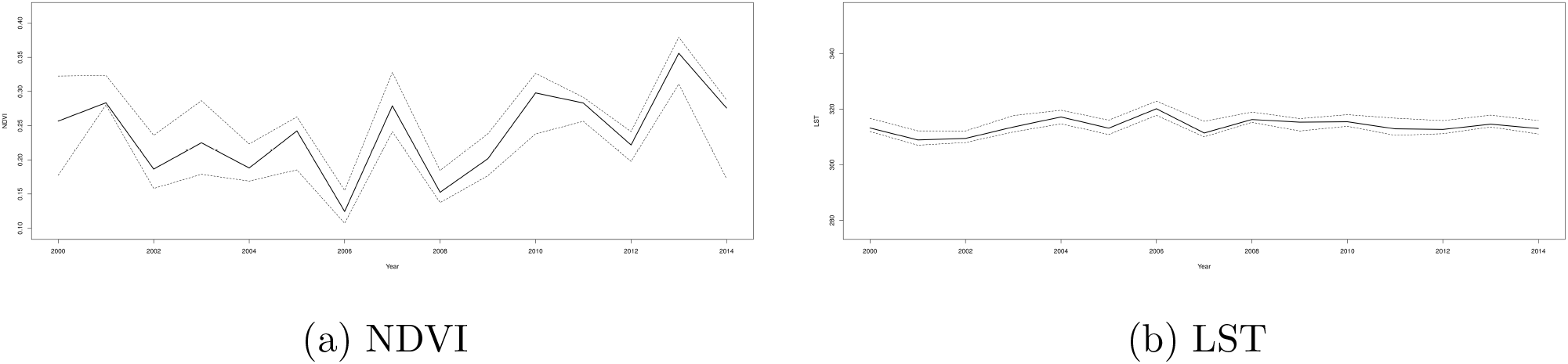
NDVI (a) and LST (b) over time in study counties, South Sudan

**Figure 4.22:**
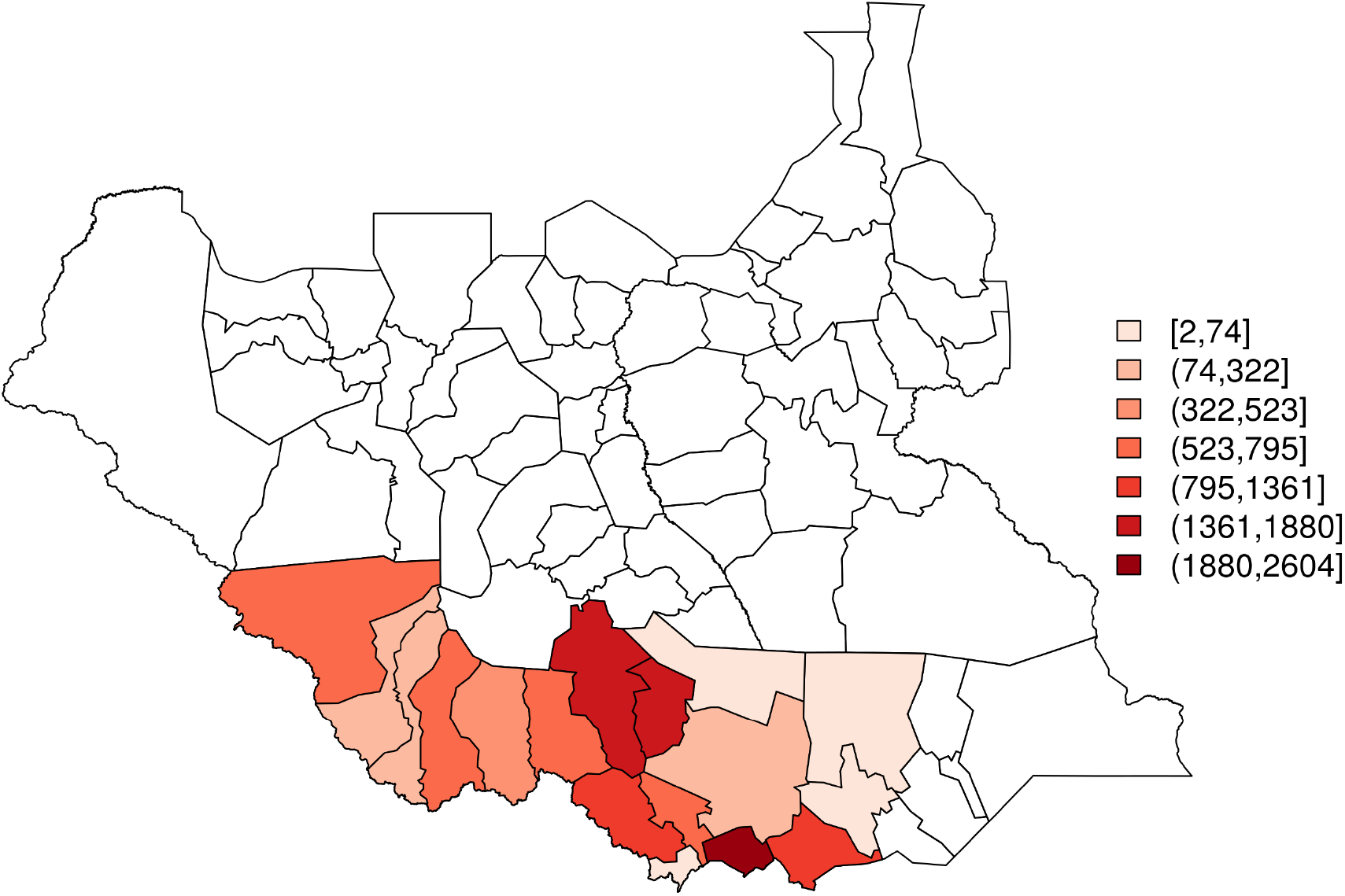
HAT cases 2000-2014, South Sudan

**Figure 4.23:**
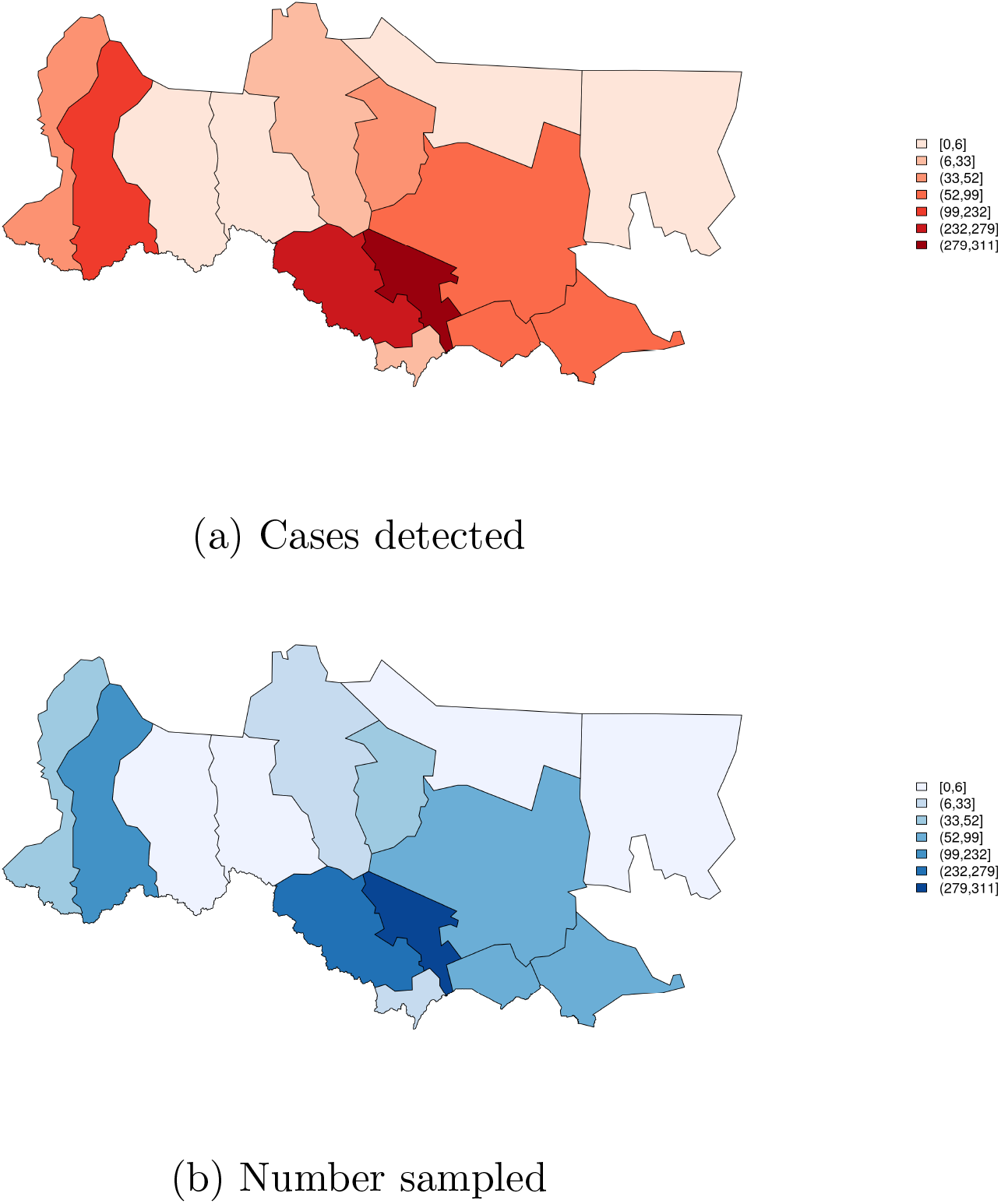
Total cases detected (a) and number sampled (b) by active surveillance in South Sudan, 2008 (restricted to active surveillance area)

**Figure 4.24:**
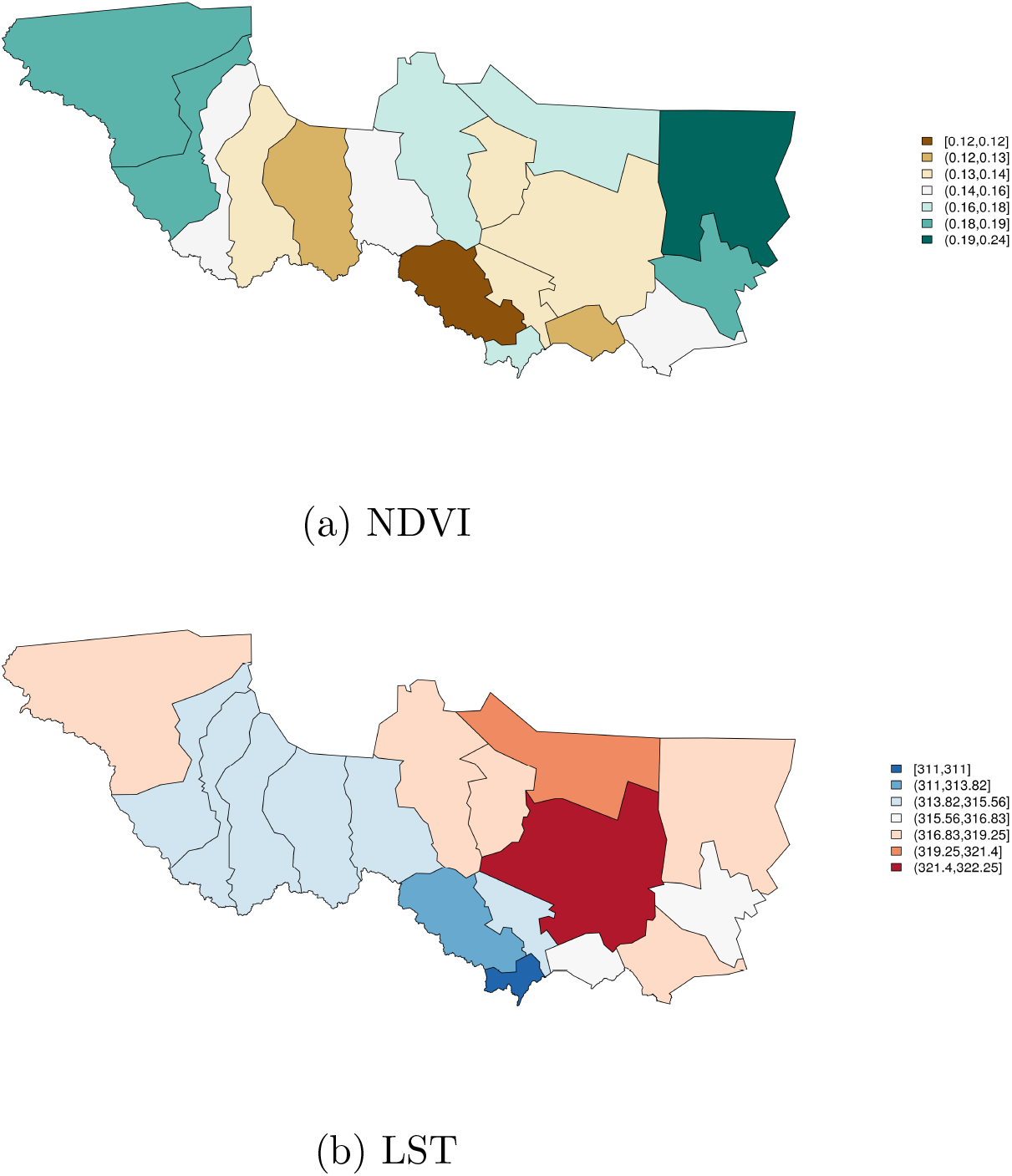
2008 NDVI (a) and LST (b), South Sudan (study area)

**Figure 4.25:**
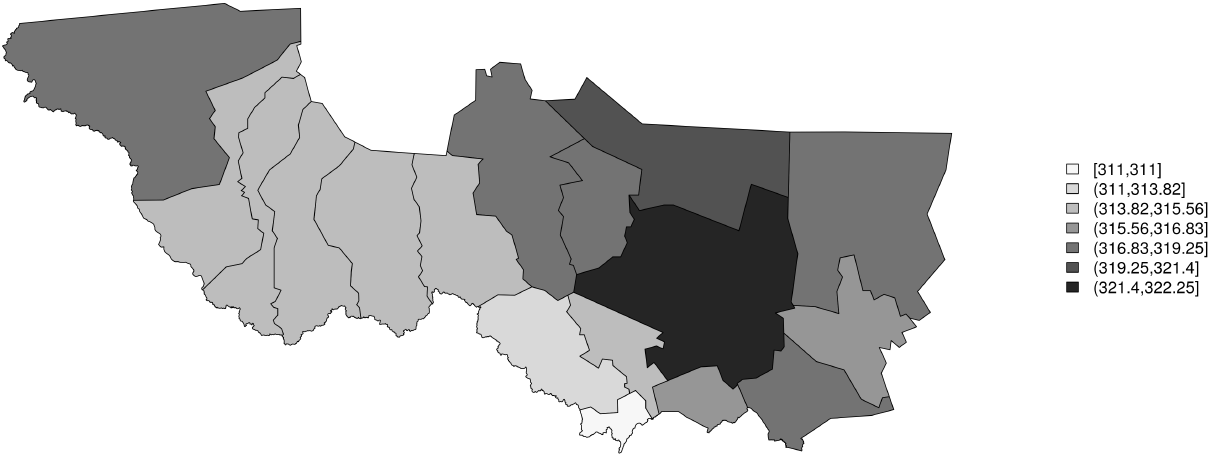
Elevation, South Sudan (study counties)

**Figure 4.26:**
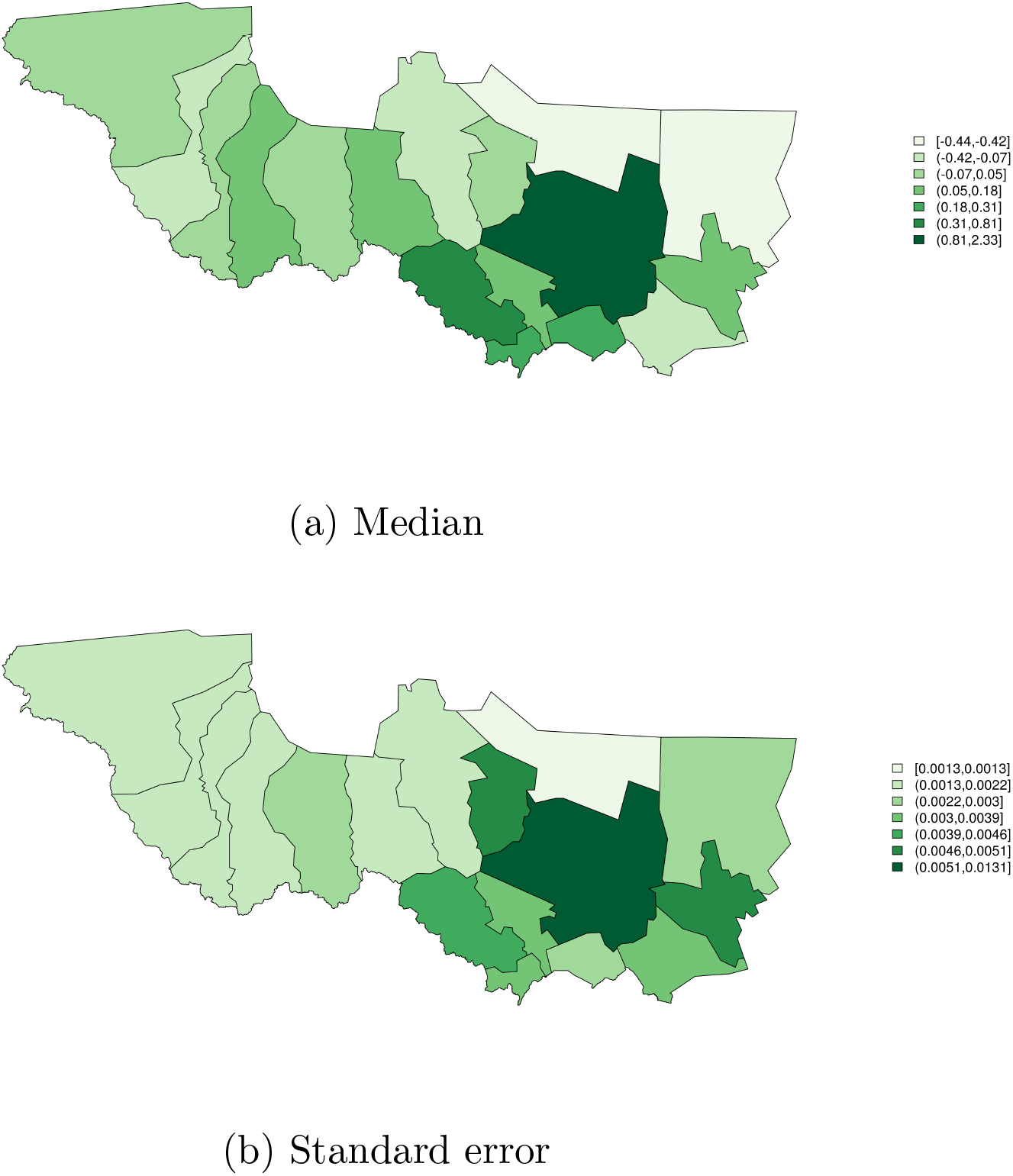
2008 wealth, median (a) and standard error (b), South Sudan (study counties)

## Notes

### Competing Interest Statement

The authors have declared no competing interest.

### Funding Statement

JM T32 ES015459-09 National Institute of Environmental Health Sciences (NIH/NIEHS) https://www.niehs.nih.gov The funders had no role in study design, data collection and analysis, decision to publish, or preparation of the manuscript.

